# Unsupervised seizure annotation and detection with neural dynamic divergence

**DOI:** 10.64898/2026.02.15.26346325

**Authors:** William K.S. Ojemann, Zhongchuan Xu, Haoer Shi, Katie Walsh, Akash Pattnaik, Nishant Sinha, Sarah Lavelle, Carlos Aguila, Ryan Gallagher, Andrew Revell, Joshua J. LaRocque, Jacob Korzun, Catherine V. Kulick-Soper, Daniel J. Zhou, Peter D. Galer, Saurabh R. Sinha, Russell T. Shinohara, Kathryn A. Davis, Brian Litt, Erin C. Conrad

## Abstract

Annotating seizure onset and spread in intracranial EEG is essential for epilepsy surgical planning, yet manual annotation is unreliable and cannot scale to large datasets. We introduce Neural Dynamic Divergence (NDD), an unsupervised framework that detects seizure activity by measuring deviation from patient-specific baseline neural dynamics using autoregressive models. NDD requires no labeled training data and adapts to individual patients, channels, and brain states. Validating against expert consensus annotations from 46 seizures, NDD achieves human-level agreement (*ϕ* = 0.58 vs. inter-rater *ϕ* = 0.64) and outperforms existing algorithms on 1,019 seizures with soft labels (AUROC = 0.87). We demonstrate clinical utility by automatically annotating 2,017 seizures, revealing that seizure spread patterns distinguish epilepsy subtypes and predict surgical outcomes. NDD also generalizes to continuous ICU scalp EEG monitoring (AUROC = 0.77). We provide NDD as an open-source Python package to enable scalable seizure annotation across research centers.

## Introduction

The propagation of neural activity facilitates communication between local and distant neuronal populations, and underlies multiple physiological, behavioral, and aberrant neural phenomena ^1^. Epilepsy is a neurological disorder characterized by the sporadic onset and spread of abnormally synchronous neural oscillations, or seizures^2,3^. Seizures can cause loss of awareness, impaired motor control, and in rare cases, death^4,5^. Tracking seizures – their occurrence, onset location, and spread – is central to epilepsy care^6,7^. Clinicians rely on this information to evaluate the effectiveness of pharmaceutical and surgical treatments and to identify seizure-generating tissue to target resection, ablation, or neurostimulation. Despite these invasive diagnostic and therapeutic treatments, many patients continue to have uncontrolled seizures^8^. Currently, the gold standard for annotating seizure onset and spread both to detect seizures and to localize seizure-generating tissue is manual annotation by expert epileptologists, an approach limited by low inter-rater reliability^9–14^. Furthermore, in intracranial EEG (iEEG) recordings, where hundreds of channels capture neural activity, manually tracking seizure spread across the implanted brain network is challenging even for a single seizure, and is impossible for large numbers of seizures and patients (Figure 1A). The growing amount of neural data collected as part of the clinical pipeline for epilepsy treatment, and the intractable scale of manual annotation suggest an urgent clinical and scientific need for an automated seizure annotation algorithm.

**Fig. 1.**
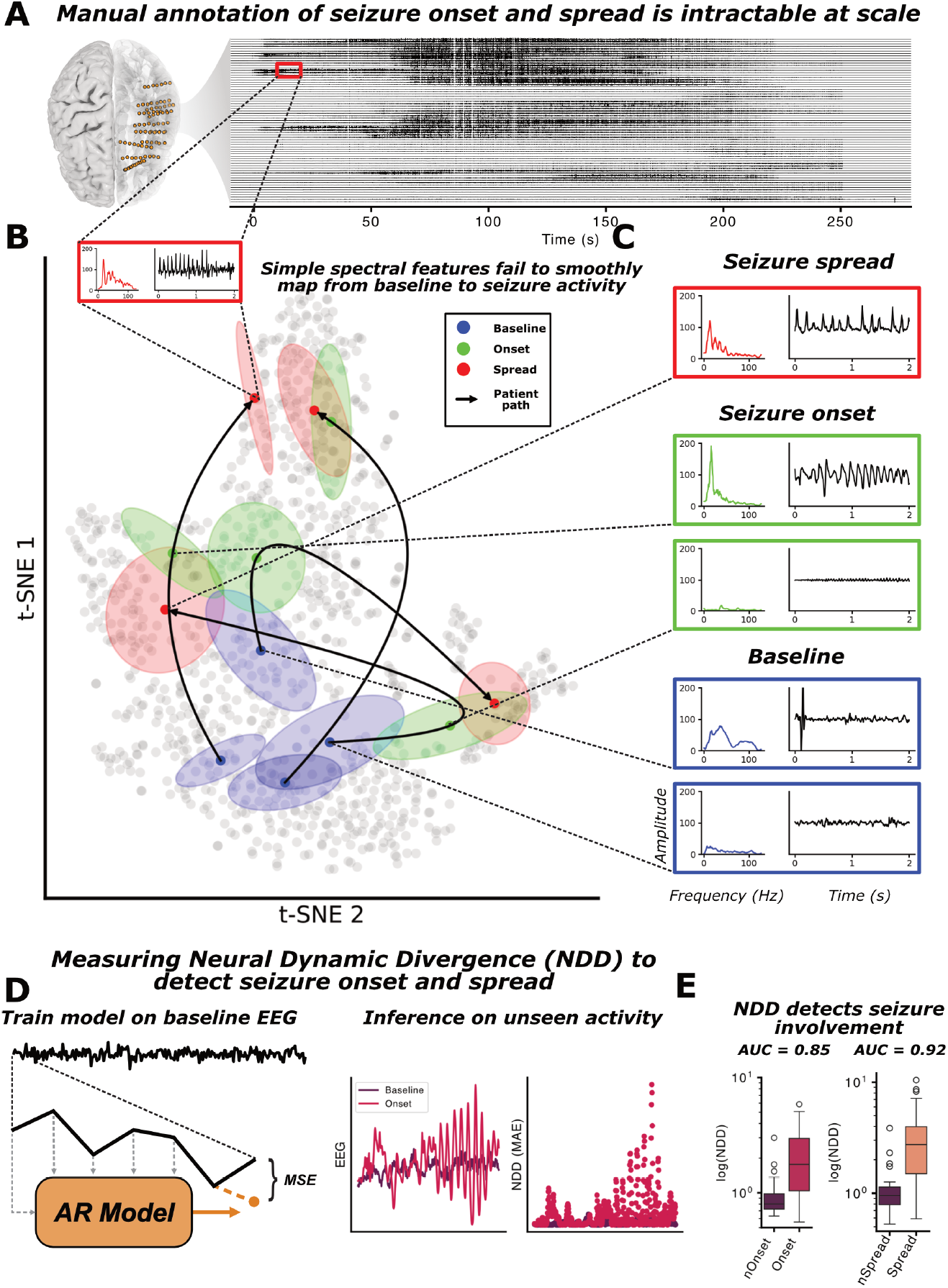
A theoretical framework for seizure onset detection: **A**) A multi-channel stereo-EEG implant and recording of the onset and propagation of an epileptic seizure, demonstrating that seizures may spread to involve many channels. **B**) A low-dimensional map of the spectral embedding of baseline (blue), seizure onset (green), and seizure spread (red) activity. Black arrows show example state transition pathways that patients take in this embedding space, highlighting the diverse and non-linear pathways that a seizure onset and spread detector needs to be sensitive to. **C**) Insets show specific spectral and time-domain examples of different seizure phases. **D**) Schematic illustrating the neural dynamic divergence (NDD) algorithm framework. An autoregressive (AR) model is trained to reproduce baseline neural dynamics. **E**) The model has high prediction loss when it encounters seizure onset (red) and spread (orange) activity, especially when compared to baseline (purple). This loss is used to detect ictal activity.

Mapping the spatial extent of multiple seizures from a single patient, or thousands of seizures across the increasingly large corpus of recorded neural activity, requires the application of quantitative algorithms. Seizure detection algorithms for intracranial EEG have rapidly progressed to the point where classifying clearly seizing and non-seizure activity—seizure detection—is almost trivial^15^. However, translating these models to precisely detect and localize seizure onset and spread— seizure annotation—is much more challenging. One impediment to solving this problem is the lack of training data with labeled seizure onset and spread. Another is the tremendous variability in seizure onset and spread activity: Seizure onset and spread patterns can vary between patients due to technical recording factors, seizure onset region, lesional status, and genetic etiology^16^. They also vary *within* patients (Figure 1B, C) due to changes in anti-seizure medication levels^17^ and transitions through different brain states on circadian and multidien time scales^18–20^. These sources of inter- and intra-patient variability manifests as a non-linear relationship between baseline, seizure onset, and spread (Figure 1B) across and within patients. This presents a significant hurdle for models designed to detect specific types of seizure onset or spread activity^21–25^.

To address the outlined challenges with annotating seizure onset and spread, and to remove the need for manual seizure annotation, we introduce neural dynamic divergence (*NDD*) and a linear approximation (*LiNDDA*)—a theoretically grounded anomaly detection algorithm well suited to mapping the transition from interictal (between seizure) to ictal (seizure) activity. The crux of the *NDD* algorithm is a multivariate autoregressive model trained on patient-, channel-, and state-specific baseline neural signals (Figure 1D). When this model is then deployed on unseen neural data, the autoregressive loss is a powerful predictor of divergence from baseline to seizure activity (Figure 1E).

To assess the accuracy of the proposed *NDD* model Figure 2 for annotating seizures recorded from intracranial EEG (iEEG), we validate it against two independent datasets of expert clinician annotations as well as publicly available benchmark algorithms (Figure 3). First, we show that *NDD* encodes ictal activity at human-level reliability, and achieves state of the art performance for seizure onset and spread detection when compared to a suite of published open source algorithms. We next demonstrate the utility of the *NDD* algorithm by annotating and detecting seizures at scale and in different modalities. We automatically annotate a large database of iEEG seizures with spatiotemporal precision and demonstrate that our algorithm supports long-held clinical hypotheses about patterns of seizure onset and spread (Figure 4). Using these annotations, we explore the potential for clinical application of our model by associating seizure spread patterns with post-surgical outcome (Figure 5). Finally, we also demonstrate the flexible nature of our algorithms by applying them to an additional EEG modality and task: detecting seizures in continuous ICU scalp EEG (Figure 6). While we primarily apply and validate our model in the context of epilepsy, the underlying framework can be applied to any pathological or evoked neural signal of interest.

**Fig. 2.**
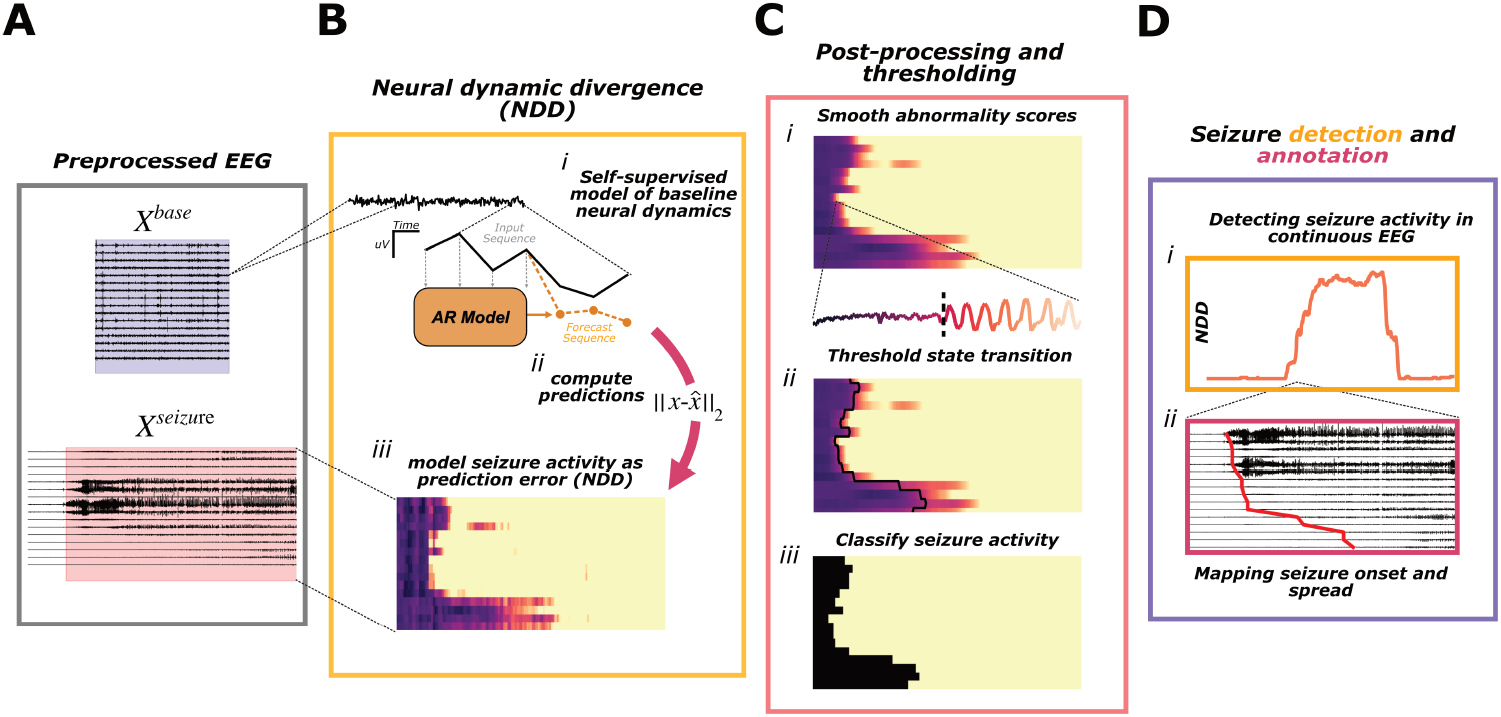
The *NDD* pipeline for seizure annotation and detection: **A**) Preprocessed baseline, *X*^*base*^, and seizure, *X*^*seizure*^, activity recorded from a single patient from intracranial EEG contacts. **B**) (i) Fitting the autoregressive (AR) model to learn the dynamics of the baseline signal on each channel, creating a generative surrogate brain model. (ii) The model is then used to generate predictions on new time series and the loss measured as MAE. (iii) This prediction error is used to measure signal abnormality that can be used to detect seizure activity. **C**) (i) Filtering the *NDD* signal to reduce the incidence of false positives. (ii) Thresholding the *NDD* signal to map the interictal to ictal state transition and (iii) classifying seizure activity with spatio-temporal granularity. **D**) (i) Using these annotations we can track seizure likelihood in continuous EEG (percent of implanted channels seizing shown), and (ii) annotate the onset and spread of seizure activity throughout the brain.

**Fig. 3.**
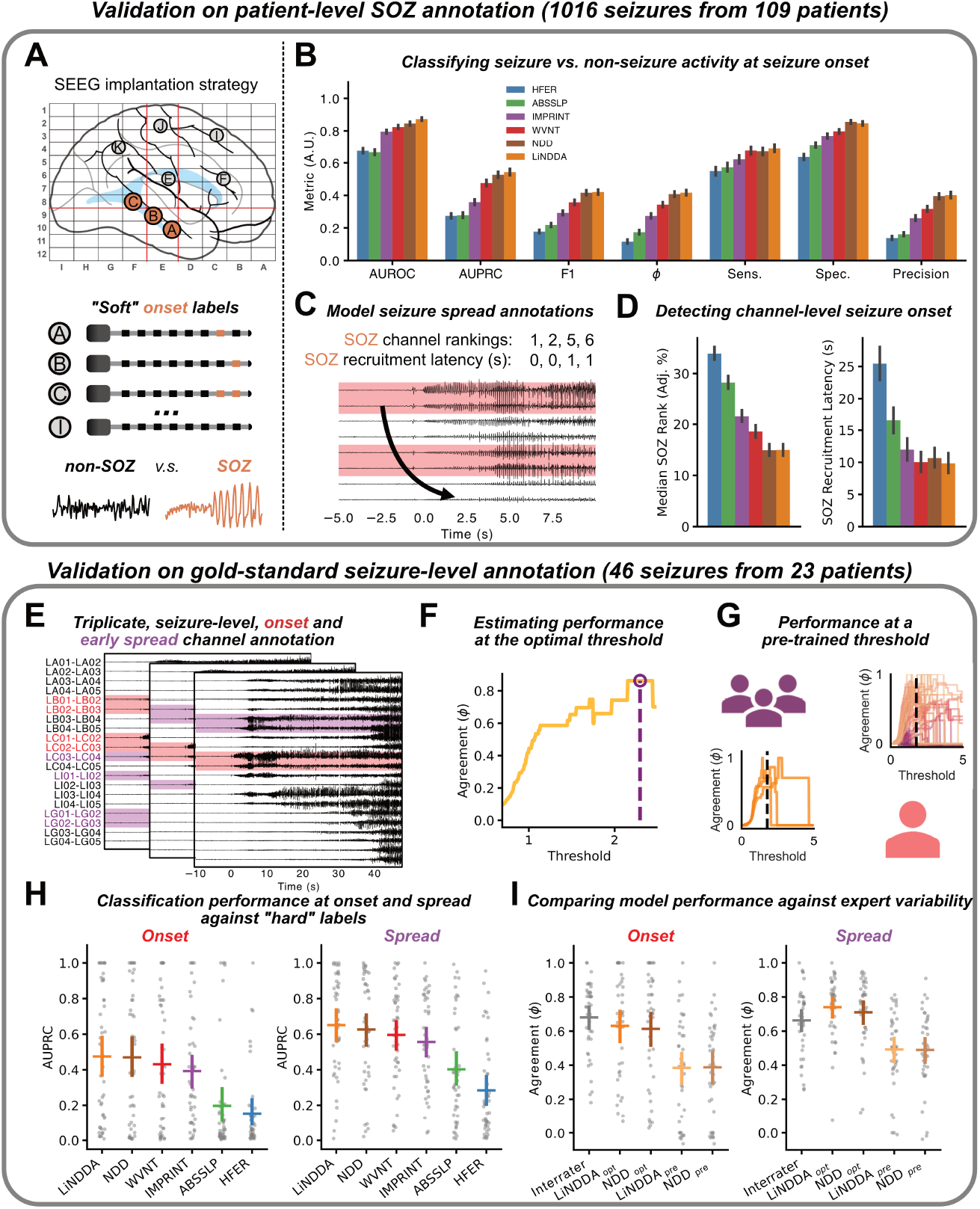
Model validation against human raters: **A**) An example stereo EEG implantation strategy where lettered depth electrodes are implanted to record intracranial neural activity from recording contacts. An expert clinical team assigned “soft” seizure onset labels at the patient level for 109 patients. We first evaluate models for their ability to discriminate between non-seizure (black) and seizure (red) activity at seizure onset, with plots showing performance across a spectrum of metrics shown in (**B**). *NDD* (brown) and *LiNNDA* (orange) had highest performance for each metric. **C**) Schematic and **D**) plots showing the spatio-temporal evaluation of model performance with SOZ channel ranking and SOZ channel onset latency. Lower-numbered rankings and shorter latencies indicate better performance. **E**) On an independent cohort we had multiple expert clinical annotators annotate seizure onset channels and channels (red) and channels seizing after 10 seconds of spread (purple). We compared *NDD* and benchmark models to the consensus annotation at two different thresholds: **F**) The optimal threshold for each seizure to assess maximum performance for each model, and **G**) at a pre-trained threshold tuned to maximize F1 score on the pre-trained dataset. **H**) Non-binary performance metrics for seizure onset activity classification (AUPRC) and (**I**) comparison to interrater reliability at different thresholds. Performance metrics and comparison statistics for all models and datasets are available in Supplementary Table 1 and Supplementary Table 2 respectively.

**Fig. 4.**
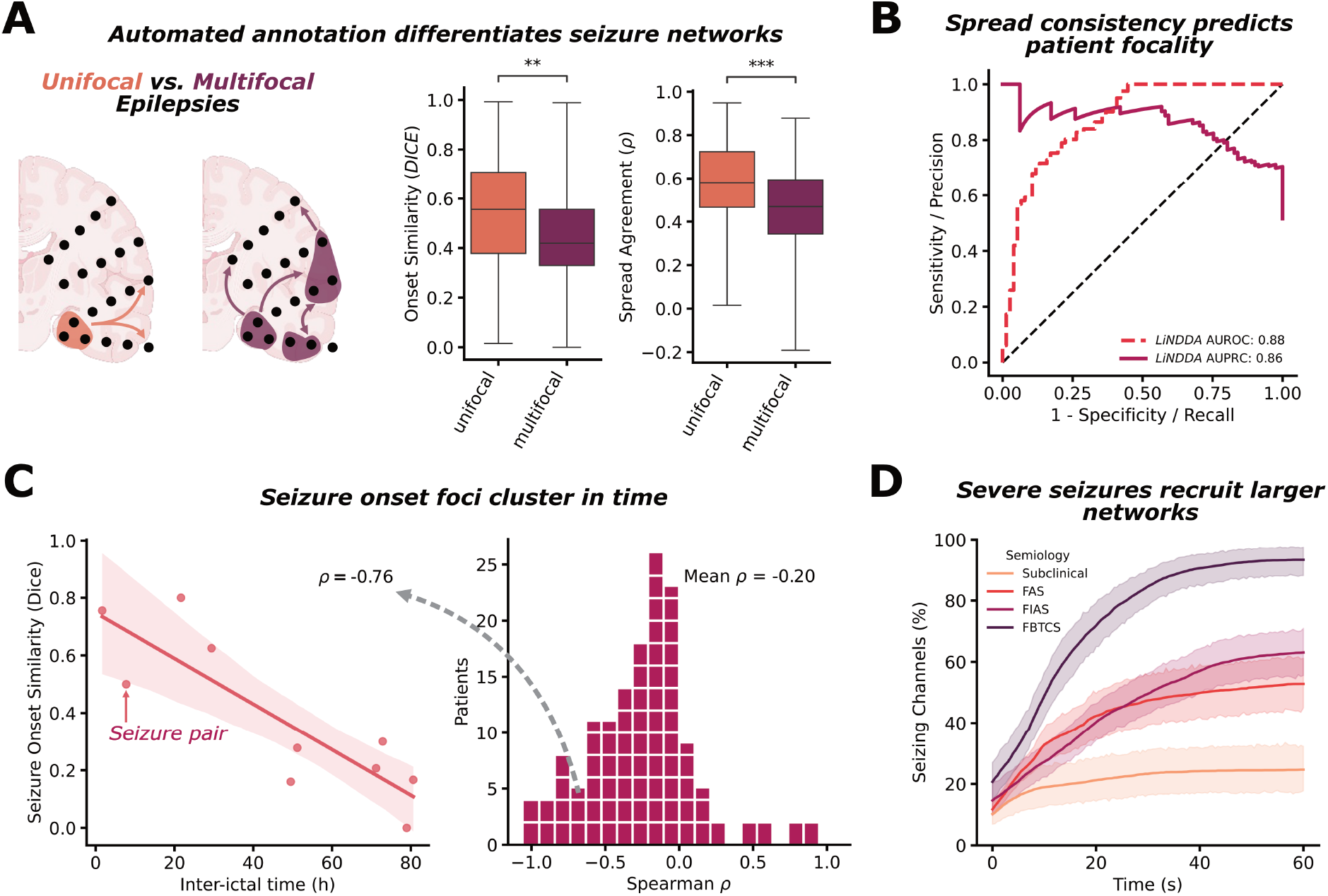
Clinical applications of. *NDD*: Demonstration of *NDD* algorithm applied to test clinical hypotheses. **A**) Seizure onset zone and spread pattern similarity within a patient distinguish unifocal (orange; more consistent patterns across seizures) from multifocal epilepsies (purple; variable onset locations across seizures). Unifocal patients show significantly higher onset similarity (Dice) and spread agreement (Spearman *ρ*) than multifocal patients. **B**) Using spread rank agreement, a logistic regression classifier predicts network focality with strong performance on unseen patients (red: AUROC = 0.88, purple: AUPRC = 0.86). **C**) Seizure onset foci cluster in time: seizures occurring closer together have more similar onset zones. Left: example patient showing strong negative correlation between inter-ictal time and onset similarity (*ρ*= −0.76). Right: distribution of within-patient correlations across the cohort (mean *ρ*= −0.20). **D**) Severe seizures recruit larger networks: the percentage of channels seizing increases with semiology severity, from subclinical seizures through focal preserved consciousness (FPC), focal impaired consciousness (FIC), to focal-to-bilateral tonic-clonic (FBTC) seizures.

**Fig. 5.**
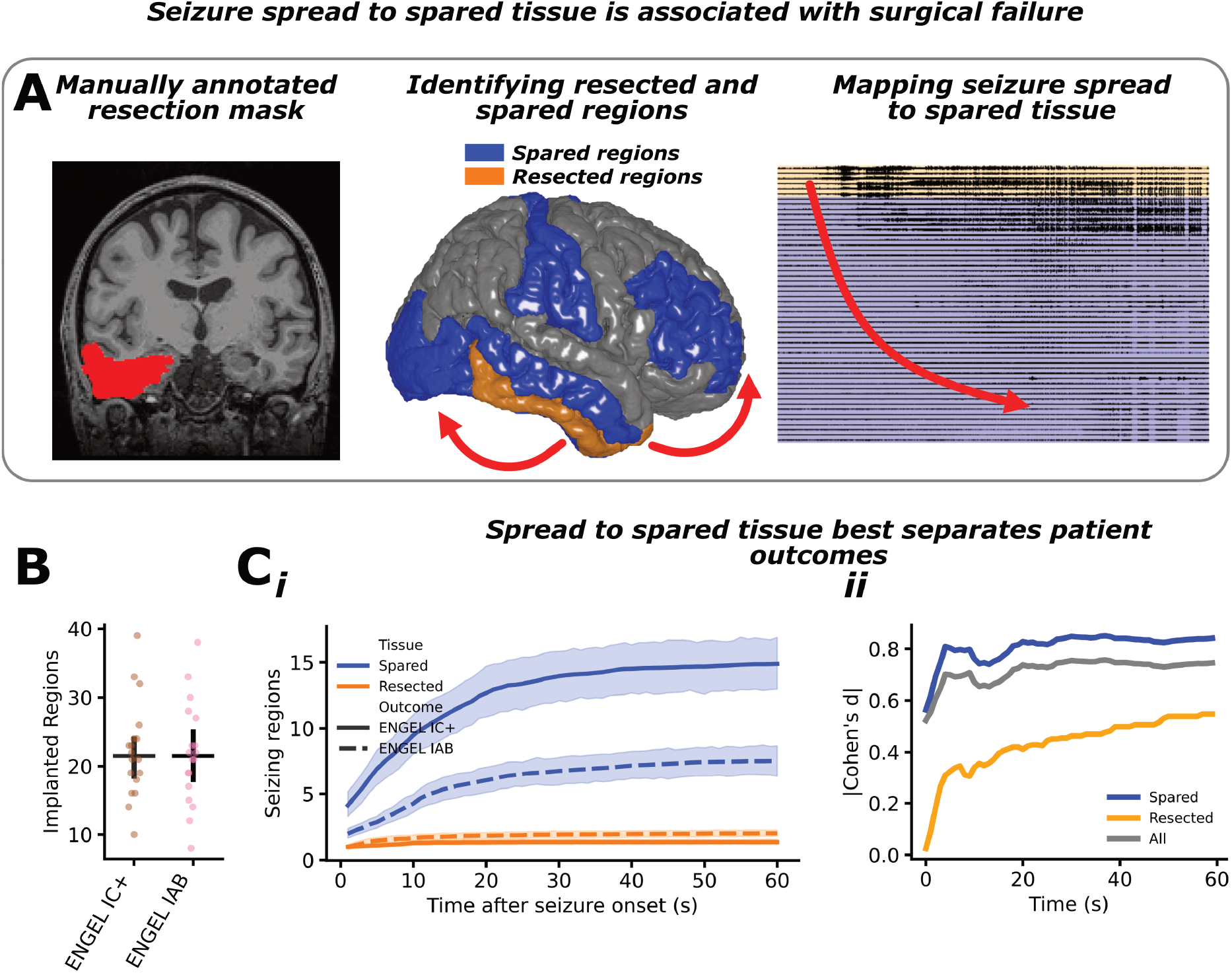
Seizure spread outside resected tissue is associated with poor surgical outcomes: **A**) We sought to examine the relationship between seizure spread and seizure freedom. Epilepsy neurologists manually annotated the resected tissue in the post-operative CT scan. We then mapped the resected tissue to regions of interest (ROI), and mapped seizure spread to these ROIs. **B**) We found no difference in the number of implanted regions by outcome, indicating that we could analyze seizure spread to regions directly. **C**) There was a non-significant trend toward higher seizure spread to spared tissue in poor outcome patients than good outcome patients at seizure onset (t = 1, mixed-effect model, *β*= 0.770, *SE* = 0.570,*p* = 0.0915, one-tailed test against the null hypothesis that poor outcome patients do not have more extensive spread) and the effect is stronger, plateauing at 5 seconds after seizure onset (t = 5, mixed-effect model, *β*= 1.381, *SE* = 0.889,*p* = 0.065, one-tailed test against the null hypothesis that poor outcome patients do not have more extensive spread).

**Fig. 6.**
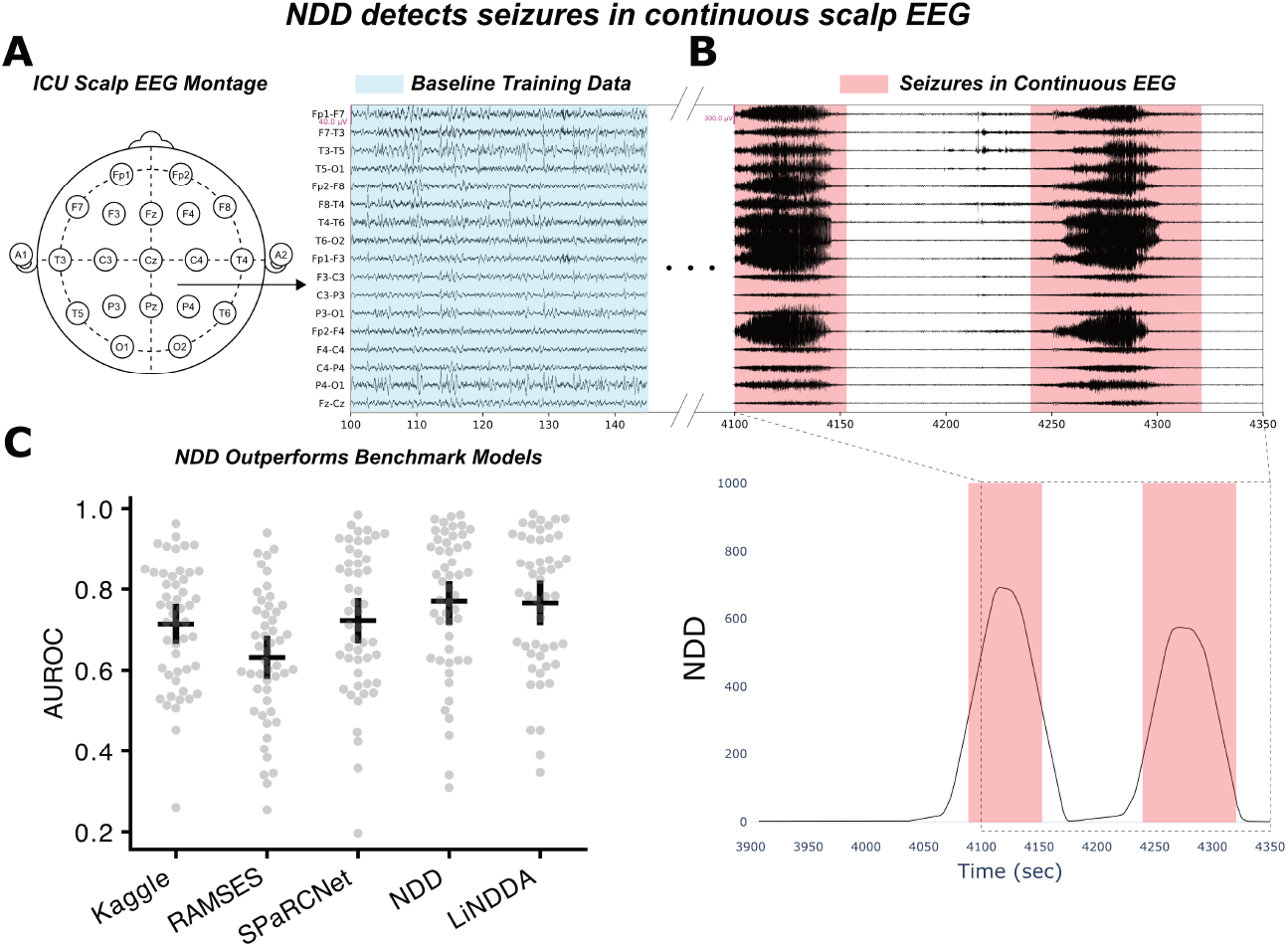
NDD generalizes to continuous seizure detection in scalp EEG: **A**) Example 10-20 EEG montage used to record neural activity from the surface of the scalp during extended, continuous EEG monitoring in the intensive care unit. The first 20 minutes of recorded activity (blue) were used to train an *NDD* model. **B**) The average NDD value across channels increased by orders of magnitude during clinician-marked seizures recorded during continuous monitoring (red). **C**) The two *NDD* models were the strongest performing seizure detectors across 50 patients when compared to state-of-the-art, open-source detection models.

One of the most pressing limitations hindering the translation of quantitative tools to improve outcomes from surgical therapies for epilepsy is their inability to scale across datasets and research centers. Towards this goal, we provide our algorithm and supporting code as a dynamic seizure detection python package, to enable rapid scaling to local and federated datasets.

## Results

### A. Neural Dynamic Divergence

Neural dynamic divergence, or *NDD*, is a framework for quantifying deviations from baseline neural activity. Conceptually, *NDD* learns a computational surrogate of the brain’s oscillatory dynamics—a generative autoregressive (AR) model—and measures changes in the underlying brain state using prediction error. First, the AR model is trained to predict the next sample of neural activity on a representative sample of baseline brain data (Figure 2A) and is then used to generate neural activity from learned dynamics (Figure 2B). As the brain transitions away from a baseline state, the AR model fails to accurately replicate the observed neural dynamics, giving a measure of abnormality (mean absolute error, MAE) that can be used to robustly detect transitions to pathological physiology (*LiNDDA* shown; Figure 1E, Figure 2). Here, we validate two different underlying AR models in the *NDD* algorithm: a recurrent neural network that predicts the next sample of neural activity through an updated latent representation, denoted simply *NDD*, and a multivariate AR (MVAR) model that forms a linear approximation of the underlying dynamics (*LiNDDA*). While we explicitly test these two models within the *NDD* framework, the algorithm is flexible and contains infrastructure to support any autoregressive architecture capable of modeling multivariate brain dynamics. Both recurrent neural networks and MVARs have been extensively studied and validated for their ability to model neural dynamical systems^26–28^. However, here we seek to validate the theoretical *NDD* framework for its ability to detect the complex and non-linear transition from baseline to seizure activity.

### B. *NDD* accurately detects clinician-annotated seizure activity at scale

We first validated *NDD* using a large cohort with non-adjudicated clinical seizure onset zone (SOZ) labels. We curated a dataset of 1019 seizures recorded on iEEG from 106 patients (average 9.6 *±* 17.3 seizures per patient) with a single suspected seizure generating focus and patient-level annotations for suspected seizure onset channels derived from the clinical reports (Figure 3A). While these labels indicate which channels the seizures are likely to come from, they are limited because 1) they are not adjudicated amongst multiple experts, and 2) they are not robust to intra-patient seizure onset zone variation. However, the massive scale of the dataset allows us to evaluate models in a real-world setting where small variations in performance may be impactful. We used these “soft-labels” to establish the performance of the *NDD* and *LiNDDA* models along with representative benchmark models for annotating seizure onset vs. non-seizure onset activity. Our benchmark models consisted of a suite of algorithms that have been proposed or validated as seizure annotation tools: high-frequency energy ratio (HFER), a measure of relative beta and gamma activity^21,29^; absolute slope (ABSSLP), a commonly used univariate seizure detection feature^17,30,31^; IMPRINT, a multivariate anomaly detection model based on a series of frequency- and time-domain features^32^; and a pre-trained WaveNet style convolutional neural network designed for single-channel seizure detection and annotation (WVNT)^30^. All performance metrics and statistics are available in Supplementary Table 1 and Supplementary Table 2 respectively.

We first evaluated our models for their ability to discriminate between seizing and non-seizing channels at the clinician defined seizure onset time. *NDD* and *LiNDDA* achieved state-of-the art performance with significantly higher AUROC (*NDD* = 0.85 *±* 0.01, *LiNDDA* = 0.87 *±* 0.01) and AUPRC (*NDD* = 0.53 *±* 0.02, *LiNDDA* = 0.54 *±* 0.02) scores than all benchmark models (Figure 3B; Supplementary Table 1, Supplementary Table 2) (see methods for a description of performance metrics). While each of these models are pretrained or self-supervised, we tested thresholded prediction metrics in an approximated leave-one-patient-out paradigm by averaging the optimal threshold for each patient across all patients (N = 106; Figure 3G). *NDD* and *LiNDDA* both continued to achieve significantly higher F1 (*NDD* = 0.42*±*0.01, *LiNDDA* = 0.42*±*0.02), *ϕ* (*NDD* = 0.41 *±* 0.02, *LiNDDA* = 0.42 *±* 0.02), specificity (*NDD* = 0.86 *±* 0.01, *LiNDDA* = 0.85 *±* 0.01), and precision (*NDD* = 0.40 *±* 0.02, *LiNDDA* = 0.40 *±* 0.02) scores than any benchmark model, while *LiNDDA* (0.69 *±* 0.02) and *NDD* (0.67 *±* 0.2) had similar sensitivity to WVNT (0.68 *±* 0.02)(Figure 3B).

We also measured the ability of each algorithm to identify onset channels in a full recording of a seizure (Figure 3C). For every seizure, Each model assigned a seizure onset time for each channel and ranked the order of seizure spread throughout the implanted network. These seizure spread annotations tested the precision—percent rank of seizure onset zone channels—and sensitivity—average latency of seizure onset zone channels—while considering the full seizure spread. Amongst models we evaluated, *NDD* and *LiNDDA* algorithms consistently ranked SOZ channels lower than any of the other algorithms (*NDD* = 15 *±* 1.1%, *LiNDDA* = 15 *±* 1.1%), suggesting superior precision in mapping the propagation of seizure activity. Further, *LiNDDA* had the lowest detected seizure onset recruitment latency of any model (9.8 ± 1.5*s*) though it was similar to WVNT (10.0 ± 1.5*s* (Figure 3D). Overall, these results indicate that the theoretical framework of neural dynamic divergence is a powerful and state-of-the-art tool for annotating seizure activity.

### C. *NDD* annotations approach human-level performance in a gold-standard annotated dataset

To perform additional validation on precise, adjudicated annotations and to compare model performance against human-level variability, we studied an independent cohort of 46 seizures from 23 patients in which trained neurologists performed triplicate seizure-level annotations of seizure onset and 10-second spread channels. (Figure 3E)^12^. We aggregated 3 or more expert annotations for seizure onset and spread channels to create two benchmarks: a gold-standard, consensus set of onset and spread channels via majority voting, and the expected level of agreement between expert annotators.

We again demonstrate that the *LiNDDA* and *NDD* algorithms have state of the art seizure annotation capabilities with the highest AUROC (*NDD* = 0.81 ± 0.08, *LiNDDA* = 0.88 ± 0.06) and AUPRC (*NDD* = 0.47 ± 0.11, *LiNDDA* = 0.47 ± 0.11) benchmarked against the consensus annotations for determining which channels were seizing at seizure onset (Figure 3H). Performance for all models was generally higher at detecting channels seizing at the 10-second spread time point than at onset, likely reflecting that ictal activity is more pronounced and homogeneous after seizure initiation (for example, *LiNDDA* AUPRC at onset = 0.47 ± 0.11 versus spread = 0.65 ± 0.08; Figure 3H,I).

We next benchmarked model performance at pre-specified thresholds against expert interrater agreement. We used Matthew’s correlation coefficient (*ϕ*) to quantify both the similarity between two expert annotators, as well as between each *NDD* model’s predictions and the consensus annotations. We generated predictions using two sets of thresholds: one optimized to each seizure (Figure 3F) and the average optimal threshold from the independent dataset (Figure 3G). At the optimal threshold, *NDD* (*ϕ* = 0.57 ± 0.10) and *LiNDDA* (*ϕ* = 0.58 ± 0.09) are capable of encoding seizure onset activity at human level performance (*ϕ* = 0.64 ± 0.07), while at a pre-trained, fixed threshold both *NDD* (*ϕ* = 0.35 ± 0.08) and *LiNDDA* (*ϕ* = 0.35 ± 0.09) performed below human-level agreement (Figure 3I). The decrease in performance from the seizure-level threshold optimization to the pre-trained, fixed threshold suggests that there is significant inter-patient and inter-seizure variance in optimal threshold limiting model performance. This motivates future work in applying unsupervised thresholding algorithms and raises the following question: despite sub-expert performance at a fixed threshold, are *NDD* annotations nevertheless scientifically and clinically useful?

To demonstrate the experimental and clinical utility of completely unsupervised seizure annotations, we deployed the *LiNNDA* algorithm with the pre-trained threshold—chosen because it demonstrated consistently superior performance compared to benchmark models—on our entire cohort of epilepsy patients from the Hospital of the University of Pennsylvania (HUP).

### D. Experimental validation on clinical and research applications

To demonstrate the generalization of the unsupervised *NDD* annotations, we next deployed the *LiNDDA* algorithm at scale on a cohort of 2017 seizures from 162 patients implanted with iEEG electrodes at HUP. For each seizure, we fit a *LiNDDA* model to the pre-ictal state (−120 s to −60 s from seizure onset) to teach the model patient-, channel-, and state-specific baseline neural dynamics. We then used the model annotations to quantify cumulative dynamic seizure spread throughout the implanted network. First, we used the channel-level spread annotations to test previously-described clinical and research hypotheses regarding seizure dynamics^17,18,33^. Then, we related seizure spread to patient outcome, demonstrating how *LiNDDA*, and by extension the broader family of *NDD* models, can be integrated into a computational system to improve surgical planning.

### E. Clinical correlates of seizure onset and spread patterns

To further validate the unsupervised *NDD* annotations, we tested whether model-defined seizure dynamics could distinguish patients based on their clinically-determined seizure onset type and based on their clinical seizure semiology.

Optimal surgical therapy depends on whether a patient’s epileptic network is unifocal—arising from a single, discrete focus amenable to resection—or multifocal—arising from multiple foci, making the patient a candidate for neuromodulation therapy (Figure 4A). We hypothesized that *NDD* would identify patients with multifocal epileptic networks as having dissimilar seizure onset and spread patterns. We compared intra-patient seizure similarity between clinically-designated unifocal (N = 100) and multifocal patients (N = 57) using two metrics: dice coefficient of onset channels and Spearman correlation of spread rank. Unifocal patients had significantly more consistent onset locations (Dice = 0.56 vs. 0.42; Mann-Whitney U(3666.5) p < 0.01) and spread patterns (*ρ* = 0.58 vs. 0.41; Mann-Whitney U(3810) p < 0.001) than multifocal patients (Figure 4A). To test whether these differences could generalize to unseen patients, we built a classifier to predict focality from spread rank similarity. The model achieved strong cross-validated performance (AUROC = 0.88, AUPRC = 0.86; Figure 4B), suggesting that *NDD*-derived seizure similarity metrics could support clinical determination of network focality.

We next examined how time between seizures, even on the scale of days in the epilepsy monitoring unit, modulates this observed seizure onset zone similarity^18,19,34^. We observed a negative correlation between inter-seizure time and seizure onset zone similarity (Figure 4C), with a mean Spearman *ρ* of −0.20 across patients, indicating that seizures occurring closer together in time arise from more similar channel sets. This is highlighted by an example patient whose seizures began arising from a new focus after 48 hours in the EMU (Dice ≈ 0.2, Spearman *ρ* = −0.76).

Seizures in patients with focal epilepsies can be classified by clinical severity (semiology): subclinical (n = 34), focal preserved consciousness (FPC) seizure (n = 51), focal impaired consciousness (FIC) seizure (n = 78), and focal-to-bilateral tonic-clonic (FBTC) seizure (n = 74)^5^. We hypothesized that seizures with more extensive spread, recruiting motor, speech, and cognitive brain regions, would in turn have more severe semiologies. We compared the percent of channels seizing at 20 seconds across semiologies and observed a significant difference at the group level (Kruskal-Wallis(67.8) p < 0.0001). Post-hoc analysis confirmed that increasingly severe semiologies had more extensive spread (Mann-Whitney U p < 0.001), with the exception of FPC and FIC seizures which showed similar spread extent (Supplementary Table 3). This demonstrates how NDD-measured seizure spread can identify clinically-severe seizures.

### F. Seizure spread outside surgical resection is associated with poor surgical outcome

To evaluate whether the patterns in seizure onset and spread identified by the *NDD* algorithm have practical implications for surgical planning, we analyzed a subset of 283 seizures from 36 patients who received resection or ablation as part of their treatment of epilepsy. Channels were mapped to the Desikan-Killiany atlas to help account for differences in the extent of implants between good (Engel IAB, n = 18) and poor (ENGEL 1C+, n = 18) outcome patients^35^. We saw no significant differences between the number of implanted regions between outcomes (Mann-Whitney U(156) p = 0.86), a common confounder of seizure spread analyses^30^. Regions were considered spared (outside the resection mask) if there were no resected channels in the region of interest (Figure 5B).

Seizure spread to spared regions trended towards separating good and poor outcome patients at seizure onset (Cohen’s d = 0.56, mixed-effect model, p = 0.09, one-tailed) (Figure 5C). There was also a trend toward greater spread outside the resection zone after 5 seconds of seizure spread in patients with poor outcomes (Cohen’s d = 0.81, mixed-effect model p = 0.06, one-tailed). Spread to spared tissue (Cohen’s d at t = 5s: 0.81) was more strongly associated with outcome than seizure spread regardless of resection status (Cohen’s d at t = 5s: 0.70).

### G. NDD detects seizures in continuous ICU scalp EEG monitoring

The hypothesis underlying *NDD* - that epileptic activity can be modeled as divergence from baseline - is not unique to intracranial EEG recordings of seizures. To demonstrate the ability of our *NDD* algorithm to generalize to other neural modalities and detection paradigms, we evaluated the algorithm’s seizure detection performance in a scalp EEG dataset comprising 50 patients who underwent ICU scalp EEG monitoring and had an electrographic seizure. We then compared the performance of the *NDD* model using the same post-processing hyperparameters as for intracranial EEG against three benchmark seizure detection models. Seizure times were annotated by clinician experts for both the previously published (n = 36)^36^ and publicly available datasets (n = 14).

The *NDD* model achieved a mean AUROC of 0.77 ± 0.05, significantly outperforming the RAMSES model (AUROC: 0.63 ± 0.05, Wilcoxon p < 0.001)—developed and optimized on the publicly available dataset and designed towards real-time seizure detection and non-seizure data reduction for ICU EEG data^36^—and the Kaggle model (AUROC: 0.71 ± 0.04, Wilcoxon p = 0.013)—the winning solution from the Seizure Detection Challenge hosted on Kaggle.com^37^—and trended higher than the SPaRCNet model (AUROC: 0.72 ± 0.05, Wilcoxon p = 0.099), which was pre-trained on 6,095 scalp EEGs from 2,711 patients^38^. All model performance metrics (Supplementary Table 4) and comparison statistics (Supplementary Table 5) are available in **Supplementary Material**.

## Discussion

Annotating seizure onset and spread with spatiotemporal precision is central to clinical care for epilepsy patients, yet manual annotation is limited by poor inter-rater reliability and cannot scale to the growing corpus of neural recordings. Here, we introduced Neural Dynamic Divergence (*NDD*), an unsupervised anomaly detection framework that learns patient-specific baseline neural dynamics and identifies seizure activity as deviation from this learned model. Validated against expert consensus annotations and benchmark algorithms across two independent datasets, *NDD* achieves state-of-the-art performance and approaches human-level agreement for seizure onset annotation.

### A. Methodological contributions

The *NDD* framework addresses a fundamental challenge in seizure annotation: the tremendous variability in seizure onset morphology across and within patients (Figure 1B). Rather than training classifiers to recognize specific ictal patterns—an approach limited by data scarcity and morphological diversity^22,24^—*NDD* reframes seizure detection as anomaly detection from a personalized baseline. This patient-, channel-, and state-specific approach enables robust detection without labeled training data.

Our validation demonstrates that *NDD* outperforms existing annotation algorithms on both soft-labeled (N = 1,019 seizures) and gold-standard consensus annotations (N = 46 seizures). Importantly, at the optimal threshold, *NDD* achieves agreement with consensus labels (*ϕ* = 0.58) comparable to inter-rater agreement between expert epileptologists (*ϕ* = 0.64). While performance decreased with a fixed, pre-trained threshold (*ϕ* = 0.35), this still exceeded benchmark models and enabled meaningful clinical analyses at scale. Interestingly, all models—including *NDD*—performed similarly to human raters for detecting spread channels, but *NDD* was uniquely able to encode seizure onset activity, highlighting the particular challenge posed by diverse seizure onset dynamotypes Figure 1^37,39^.

Notably, the linear *LiNDDA* approximation consistently matched or outperformed the nonlinear RNN architecture across our validation experiments. This finding aligns with recent work demonstrating that neural dynamics at macroscopic recording scales are well-approximated by linear systems^40^, a property that has enabled successful translational applications in epilepsy and broader neuroscience^28,41^. The recursive prediction strategy employed by *LiNDDA*—generating multiple time steps ahead—may further enhance sensitivity by more thoroughly interrogating the learned dynamics, making the model responsive to the broad spectrum of state transitions observed at seizure onset (Figure 1B,C)^42^. The one exception was scalp EEG, where *LiNDDA* showed slightly decreased performance relative to *NDD*. This may reflect modality-specific differences: scalp EEG measures neural activity at a coarser spatial scale with attenuated high-frequency content^43^, potentially favoring the nonlinear model’s capacity to capture subtler shifts in signal power. For intracranial EEG applications, where recordings contain rich information across the frequency spectrum, *LiNDDA* offers a clear practical advantage: comparable or superior performance with reduced computational complexity and fewer hyperparameters to tune.

### B. Clinical and scientific implications

Deploying *NDD* annotations across 2,017 seizures revealed clinically meaningful patterns that would be intractable to quantify through manual annotation. *NDD* captured the expected differences between unifocal and multifocal patients, with multifocal patients showing lower intra-patient similarity in seizure onset and spread patterns. A classifier trained on *NDD*-derived seizure similarity predicted network focality with strong performance (AUROC = 0.88), substantially exceeding reported pre-implant clinical predictors (AUROC = 0.67)^44^ and interictal EEG features (AUROC = 0.79)^33^. This distinction has direct therapeutic implications: patients with unifocal networks are stronger candidates for resective surgery, while those with multifocal networks may benefit more from neuromodulation^45^. *NDD* thus provides a validated, quantitative tool for guiding this critical treatment decision.

We also observed that seizure onset zones shift over the timescale of EMU monitoring, with seizures closer in time arising from more similar channel sets. This temporal structure has been described using high-dimensional latent variable models^18,34^, but *NDD* quantifies this phenomenon using a clinically interpretable metric: onset channel overlap. Beyond these novel findings, the ability of unsupervised *NDD* annotations to replicate phenomena recognized qualitatively by the epilepsy community—while providing robust empirical quantification—validates the clinical and scientific utility of the algorithm.

The relationship between seizure spread and clinical severity (semiology) was also encoded in *NDD* annotations: increasingly severe seizure types recruited larger portions of the implanted network, consistent with prior work^17^. Notably, focal aware and focal impaired aware seizures showed similar spread extent, suggesting that loss of awareness may depend on spread to specific brain regions rather than overall spread magnitude. This finding motivates future work examining spread to regions implicated in ictal unconsciousness^46,47^. From a translational standpoint, the ability to quickly identify seizures progressing toward bilateral tonic-clonic activity—the most dangerous seizure type, associated with falls, injury, and sudden unexpected death in epilepsy (SUDEP)^4^—would enable rapid intervention via emergency stimulation or medication therapy.

### C. Implications for surgical planning

The association between seizure spread to non-resected tissue and poor surgical outcome has important implications for clinical practice. Notably, recent work found that the proportion of seizure onset regions resected did not correlate with surgical outcome^48^, suggesting that seizure onset localization alone may be insufficient for predicting surgical success. Our findings complement this observation: the rapid increase in effect size within the first 5 seconds of seizure spread—aligning with prior reports on early recruitment of epileptogenic tissue^49–52^—indicates that spread dynamics may capture prognostically relevant information beyond what is encoded at seizure onset. Critically, these early spread patterns may explain why some patients experience seizure recurrence despite complete resection of the clinically-identified seizure onset zone.

These findings suggest that automated seizure annotation could identify tissue requiring prophylactic treatment or secondary monitoring beyond the primary seizure onset zone. Spread annotations could flag candidate regions for extended resection when seizures consistently propagate outside the planned surgical margin, or guide placement of responsive neurostimulation devices to intercept propagation when resection is not feasible. While these results are hypothesis-generating given the trending statistical significance (p = 0.06), the large effect size (Cohen’s d = 0.81) and alignment with prior literature motivate prospective validation of *NDD*-derived spread biomarkers for surgical planning.

While we primarily demonstrate the capabilities of the algorithm for performing clinical research at scale, automated seizure annotation could also impact care for individual patients. Manual annotation and visualization of seizure onset and spread is notoriously unreliable^12^, labor-intensive (Figure 1A), and a bottleneck for surgical planning^53^. Automated annotation methods like *NDD* can support EEG interpretation and surgical planning when integrated with automated imaging^54^ and visualization software (Figure S3)^55,56^.

### D. Generalization across modalities

The *NDD* algorithm also generalized to continuous ICU scalp EEG monitoring, outperforming models specifically developed for this task including RAMSES^36^ and the Kaggle competition winner^37^. This cross-modality performance demonstrates that the core hypothesis—epileptic activity as divergence from baseline dynamics—applies broadly across recording paradigms.

This generalization addresses a critical bottleneck in epilepsy care. Continuous EEG monitoring is essential for detecting non-convulsive seizures, which occur in up to 20% of critically ill patients and are associated with worse outcomes when untreated^57^. However, the volume of data generated—often days of continuous recording per patient—far exceeds the capacity of clinical neurophysiology teams. Current practice relies on periodic review or threshold-based alerts with high false-positive rates, leading to alarm fatigue and missed events. An unsupervised algorithm that learns patient- and state-specific baseline dynamics is well-suited to this setting, where inter-patient variability due to sedation, encephalopathy, or underlying pathology limits the utility of models trained on normative data. By automating initial annotation, *NDD* could serve as a triage tool—flagging high-priority segments for expert review and enabling clinicians to focus on interpretation rather than detection. The open-source availability of *NDD* lowers adoption barriers, allowing institutions to integrate automated annotation without proprietary licensing constraints.

### E. Building upon prior work

Recent advances in deep learning have produced powerful seizure detection^15^ and annotation^58^ models, including foundation models trained on large EEG corpora^59^. However, these approaches rarely address seizure onset annotation due to the sparsity of labeled training data. Unsupervised approaches—including entropy-based methods^60,61^, reconstruction loss^62^, and autoregressive models^63^—offer an alternative, though validation against precise onset annotations has been limited. Hypothesis-driven algorithms based on specific spectral signatures^21,64^ have been validated for localizing epileptogenic tissue, but are often tailored to specific onset morphologies that may not generalize across patients^24^. By validating *NDD* against both expert consensus and existing algorithms on multiple datasets, and demonstrating clinical utility at scale, we establish a benchmark for seizure annotation performance and provide annotated data to support future algorithm development^12,65^.

### F. Limitations and future directions

Several methodological considerations apply when deploying *NDD*. First, we validate seizure onset zone annotation rather than epileptogenic zone localization, which have distinct clinical meanings. Second, hyperparameters—particularly post-processing filters and detection thresholds—impact sensitivity, and optimal settings may vary across applications. The sub-optimal fixed threshold limits real-time deployment, though adaptive thresholding algorithms^66^ could address this limitation. Third, our validation uses data from a single center (HUP); multi-center validation is needed to establish generalizability across institutions and recording systems. Finally, while we examine performance across epilepsy subtypes, systematic characterization of failure modes—which seizure types or onset morphologies challenge *NDD*—remains an area for future work.

To enable broad application, we provide *NDD* as an open-source Python package (link available upon publication) that includes the full signal processing pipeline and implementations of benchmark algorithms. By lowering barriers to automated seizure annotation, we aim to facilitate both large-scale clinical research and integration into surgical planning workflows.

## Data Availability

All data analyzed are available at http://ieeg.org and on pennsieve.io at https://doi.org/10.26275/N1SW-DYMC

## Acknowledgments

We thank Carolyn Wilkinson, and all other members and staff of the Center for Neuroengineering and Therapeutics for their continued help and support in this work. We also thank John Magnotti and Zhengjia Wang for their help with data visualization.

## Funding

### Author Contributions

Conception and design of the study included WO, ZC, HS, EC, BL. Acquisition and analysis of data included WO, AP, SL, NS, RG, JL, JK, CK, DZ, CA, EM, SS, KD, BL, and EC. Software development was carried out by WO, HS, KW, ZC, and PG. Drafting a significant portion of the initial manuscript or figures included WO, ZC, HS, BL, and EC. All authors reviewed and edited the final version of the manuscript.

## Conflicts of Interest

The authors declare no competing interests.

## Materials and Methods

### A. Neural dynamic divergence

To capture deviations from typical brain activity at seizure onset and during propagation, we developed an anomaly detection approach based on using a multivariate autoregressive model, *f* (·), to simulate baseline neural dynamics. The model takes as input a sequence of observations *X*_*t*−*s*:*t*−1_ = [***x***_*t*−*s*_, ***x***_*t*−*s*+1_,…, ***x***_*t*−1_] ∈ ℝ^*m*×*s*^ to predict the next state 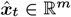, where ***x***_*t*_ is the brain state measured from *m* channels at time *t*, and *s* is the number of time step inputs. In general, the brain’s baseline neural dynamics are modeled as

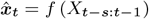

In this manuscript we test two different classes of autoregressive models; however, within the DynaSD software package, we provide infrastructure to rapidly develop new models.

The first autoregressive model, denoted simply *NDD*, is a non-linear long short-term memory (LSTM) model that processes the input sequence, updating a hidden state that is then linearly transformed to make predictions about future brain states

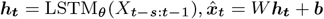

The second model is a linear vector autoregressive (VAR) framework, denoted *LiNDDA* in the manuscript, where predictions are generated as a linear combination of past observations:

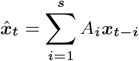

where *A*_*i*_ ∈ ℝ^*m*×*m*^ are coefficient matrices learned for each input timestep, *i*, and the constant term is set to zero due to signal normalization. The order of the model, *s* was tuned to 3 on our validation dataset of soft labels to maximize AUROC and AUPRC (allowing us to select the optimal model hyperparameters before applying any thresholding; Figure S1).

#### A.1. Learning baseline neural dynamics

To teach the model baseline, interictal dynamics on a patient, channel, and state-specific scale contained in a baseline clip of neural data, *X*^*base*^ ∈ ℝ^*m*×*L*^ ⊂*X* (Figure 2A), we train the model, *f* (·), with parameters *θ* to minimize the squared error between the predicted and observed neural activity where the optimal set of parameters

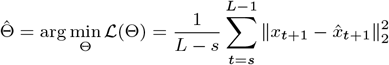

(Figure 2B).

#### A.2. Measuring divergence from baseline dynamics

Once trained, the model is applied to unseen neural signals, *X*_*sz*_ = *X\X*_*base*_, from the same patient to quantify divergence from baseline dynamics (Figure 2B). For each input sequence, *s*, we recursively generate *u* predicted samples with a stride of *u*—we only generate one predicted value of each recorded sample. For the *NDD* implementation, we set *u* = 1, while we tuned the value of *u* = 2 as part of our hyperparameter tuning for *LiNDDA* (**Supplementary Material**). We then segment *X*^*sz*^ into overlapping windows of length *w*_*l*_ = 1 s and stride *w*_*s*_ = 0.5 s where 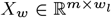, and average absolute difference between predicted and true states of recorded neural activity. Iterating over all windows yields a continuous, multi-variate time-series measuring divergence from baseline dynamics that forms the basis for seizure detection (Figure 2B).

### B. Benchmark models

We test the *NDD* algorithm against a series of representative state-of-the-art benchmark seizure annotation features and models.

#### B.1. Absolute Slope

Absolute slope (ABSSLP) has long been used to measure ictal activity, and is a correlate of line length - another low-cost seizure detection feature^67^. Absolute slope is defined on a discretely sampled single channel of neural activity, *X*_*i*_, over a period of time, *w*_*l*_, as

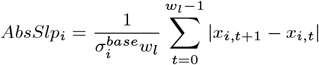

where 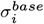 is the standard deviation of signal amplitude on channel *i* during a baseline reference period, 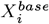.

#### B.2. High Frequency Energy Ratio

High Frequency Energy Ratio (HFER) is the feature underlying the epileptogenicity index^21^ that is calculated as the ratio between beta and gamma (12.4 - 96 Hz), and theta and alpha (3.5 - 12.4 Hz) spectral power in a given window of neural activity, *w*_*l*_.

#### B.3 IMPRINT

The IMPRINT algorithm, similar to *NDD*, measures deviation from a baseline period of activity. Rather than fitting a surrogate brain model to baseline dynamics, IMPRINT learns a multivariate distribution of features over a baseline period and measures the abnormality of new neural activity using Mahalanobis distance. The features composing this algorithm are described in detail in a previous publication^32^.

#### B.4 WaveNet

The WaveNet algorithm (WVNT) was built on a WaveNet-style architecture of causal, dilating convolutional layers and trained to discriminate between clearly seizing and clearly non-seizing neural activity. The model operates on the hypothesis that the transition from interictal to ictal activity would map in probability space between these two classes^30^. We note that WVNT was trained on a subset of activity from 13 patients that are contained in the soft-label validation dataset, which may skew estimated performance higher. However, the set of gold-standard annotations is independent from model training.

### C. Intracranial EEG data collection

We retrospectively analyzed a multi-center dataset consisting of 2017 seizures from 162 patients from the Hospital of the University of Pennsylvania (HUP) implanted with intracranial EEG electrodes as part of presurgical evaluation for drug resistant epilepsy. EEG recordings were collected at 1024 Hz from 2011 to 2024 using a Natus Quantum LTM amplifier (Natus Medical, Pleasanton, CA) The study protocol, data collection, and retrospective analysis was approved by the Institutional Review Board (IRB) of University of Pennsylvania. All EEG recordings are made available on http://ieeg.org^68^. There were two subsets of this cohort that received expert clinician annotation of the seizure onset zone. First, 1019 seizures from 106 patients were determined to be unifocal by clinical impression of their spontaneous seizures, and a multi-disciplinary care team determined a set of likely seizure generating channels as part of routine clinical care. These soft-labels, while not annotating at the seizure level, were likely to be good labels for the seizure onset zone. Second, in a post-implant analysis, 5 expert epileptologist annotators (C.K., D.Z., E.C., J.K., J.L.) performed triplicate seizure-level annotations of 46 seizures from 23 patients^12^. To rigorously validate the ability of the *NDD* model to encode and detect seizure activity throughout the different phases of seizure dynamics, seizures were annotated for channels seizing in the one second after seizure onset as well as the one second period after 10 seconds of seizure activity, or early spread.

### D. Intracranial EEG processing

We analyzed each of the 2017 seizures with a 180 second pre-ictal and 120 second post-ictal buffer sampled at 256 Hz. For seizures from patients with available electrode co-registration, electrodes assigned parcels from the Desikan-Killiany atlas parcellation using ieeg-recon^54^ and any channels in CSF or outside the head were removed from further analysis. For all seizures, we then bipolar re-referenced the signal, rejected artificially noisy channels using an automated pipeline, notch filtered at 60 and 120 Hz, bandpass filtered between 1 Hz and 120 Hz, and pre-whitened the signal. After preprocessing, the buffered seizure signal on each channel was independently scaled to a baseline inter-ictal time window from −180 s to −120 s prior to seizure onset, *X*^*base*^ using the Robust Scalar algorithm^69^. When detecting seizure activity with WVNT, we modified the preprocessing steps to align with the filtering used during model training^30^.

### E. Seizure annotation

For each seizure in the manually annotated patient cohort, we fit the *NDD* model to the normalized and preprocessed *X*_*base*_ signal, and then generated *NDD* or benchmark model values in bins of *w*_*l*_ = 1 with *w*_*s*_ = 0.5 seconds of stride. We convolved the signal with a 20 sample moving average filter to reduce the effect of spurious detections. We then applied a threshold to the *NDD* values to map the interictal to ictal transition and classify seizure activity (Figure 2C). We further defined a 1 second window as seizing if 4 of the 5 following seconds of detections were positive detections after filtering. Channels were considered seizing at onset as channels with positive seizure detections in the first second after the clinician determined onset time. Similarly, we considered early spread as channels with detections starting from 10 to 11 seconds after the seizure onset time. In addition to identifying seizing channels relative to a fixed time, we also extracted when the model first detects seizure activity on each channel after filtering (Figure 2D).

### F. Optimal Threshold Selection

We determined the optimal classification threshold on each seizure using a plateau-detection algorithm designed to identify stable regions of peak performance. For each model, we evaluated 301 equally-spaced threshold candidates between the 5th and 99th percentiles of predicted probabilities. At each threshold *τ*, we calculated the Matthew’s Correlation Coefficient and identified the maximum *ϕ* per seizure to quantify how well each model can encode seizure activity compared to interrater agreement (Figure 3F). We then computed the F1 score for all *τ*and identified all thresholds yielding performance within *ϵ*= 0.01 of the maximum F1 score. Among these near-optimal thresholds, we identified contiguous segments of consecutive thresholds and selected the midpoint of the longest segment as the optimal threshold. This approach favors thresholds that maintain high performance across a range of values, improving robustness compared to selecting the single threshold that maximizes F1 score. The pre-trained threshold from the validation dataset was generated by averaging the threshold that maximized F1 score within patients, and taking the median value across patients to avoid influence from outliers (Figure 3G).

### G. Evaluating model performance

We use a comprehensive suite of performance metrics to assess the capability of different algorithms to annotate seizure onset and spread. Here we define true and false positives, and true and false negatives as TP, FP, TN, and FN respectively.

#### G.1. Sensitivity

The percent of SOZ channels detected.

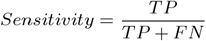

#### G.2. Specificity

The percent of non-SOZ channels correctly detected.

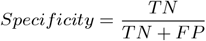

#### G.3. Precision

The percent of predicted SOZ channels that belong to the SOZ. This is equivalent to he positive predictive value.

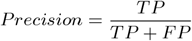

#### G.4. AUROC

The area under the receiver operator curve, or the integral of the tradeoff between false positives rate and true positive rate at different thresholds.

#### G.5. AUPRC

The area under the precision-recall curve, or the integral of the tradeoff between precision and recall (sensitivity) and precision.

#### G.6. F1 score

The geometric mean of the precision and recall.

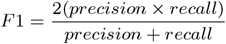

#### G.7. Matthew’s Correlation Coefficient (ϕ)

The Pearson correlation of the true labels and the predicted labels. This performance metric is suggested to be the most robust for evaluating the performance of binary predictors^70,71^.

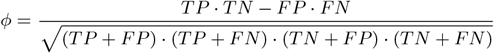

#### G.8. Adjusted median SOZ rank

We ranked channels by model-detected seizure onset time, assigning tied last place to channels with no detected activity. The adjusted median SOZ rank was calculated as the median rank of clinically-determined SOZ channels (*R*_SOZ_), adjusted by subtracting half the SOZ size (|*S*|) (yielding 0 for perfect predictions), then normalized by the total number of implanted channels (*N*_total_) and expressed as a percentage.

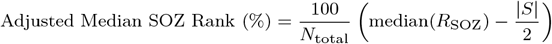

Any SOZ channels that were not detected by a given model were assigned the last possible rank.

#### G.9. Mean SOZ recruitment latency

We calculated the mean recruitment latency of the set of clinically-determined SOZ channels (*S*) as the average time from seizure onset to first detection by the model (*L*_*i*_). Channels in *S* that were not detected by the model were assigned a latency equal to the seizure duration.

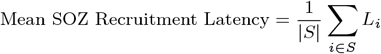

Any SOZ channels that were not detected by a given model were assigned an onset latency of the duration of the seizure.

#### G.10 Expert annotator interrater reliability

To benchmark model performance against human-level accuracy, we leveraged multiple expert annotators per seizure in our second evaluation dataset. We calculated expected human agreement by averaging the *ϕ*coefficient between each pair of annotators for a given seizure^12^. We use *ϕ*to measure annotator agreement based on its robustness to imbalanced classes^72^. We then compared this to the *ϕ*coefficient between the model’s annotations and the majority-voting consensus of the clinicians for the same seizure. We consider a model to be at human level performance if we fail to detect a significant difference^73^ from the interrater agreement.

### H. Experimental analysis of seizure spread

For each of 2017 seizures in our iEEG dataset, we annotated seizure onset times in each channel using the *LiNDDA* algorithm and converted these annotations into metrics for association with patient- and seizure-level variables.

To distinguish unifocal from multifocal epilepsies, we quantified intra-patient seizure similarity using the dice coefficient of onset channel sets (channels seizing within 1 second) and Spearman correlation of spread rank order, with non-seizing channels assigned last-place rank. We used spread rank correlation as input to a logistic regression classifier with balanced class weighting, evaluated in a leave-one-patient-out paradigm. We also examined whether seizure onset zones shift over time by correlating inter-seizure interval with onset-zone similarity.

To examine the relationship between seizure spread and semiology, we compared the proportion of channels seizing in 1 second windows a stride of 0.5 seconds across semiology categories (subclinical, focal aware, focal impaired awareness, and focal to bilateral tonic-clonic).

### I. Resection masks and electrode localization

Each of the 36 resection or ablation masks were drawn using ITK-Snap^74^ by a board-certified epileptologist (DZ) from post-surgical T1-weighted MRI scans. We then labeled channels as resected or spared by whether the nearest voxel to the electrode contact centroid. Then, using the ieeg-recon^54^ we mapped the electrodes to the Dessikan-Killiany atlas^35^ for cortical and subcortical parcellation. We considered a region to be labeled ‘resected’ if any of the channels implanted in that region were resected. We then mapped automatically detected seizure spread latency to channels, assigning the latency to the first contact of the bipolar pair. Each implanted region was assigned the minimum seizure latency of all channels in that region.

### J. Neuro ICU EEG data collection

The ICU EEG dataset consists of 50 patients admitted in ICUs at University of Pennsylvania who underwent continuous EEG monitoring between 2017 and 2024. The EEG data were recorded at 256 Hz using Natus Xltek equipment (Natus Medical, Pleasanton, CA). The electrodes were placed according to the international 10–20 system. The seizures were annotated by board-certified clinical neurophysiologists^36^. The data collection and retrospective analysis was approved by the IRB of University of Pennsylvania. All EEG recordings and annotations are available on http://ieeg.org^68^.

### K. Neuro ICU EEG processing and seizure detection

For the *NDD* model, EEG data were bandpass filtered between 1 and 40 Hz, notch filtered at 60 Hz, and bipolar re-referenced. The continuous recordings were segmented into 1-second clips with 0.5 second overlap. We trained patient-specific *NDD* and *LiNDDA* models using the first 10 minutes of interictal data as *X*^*base*^. During evaluation, we then applied the same procedure as with intracranial EEG clips to the entirety of the patients continuous ICU EEG recording, *X*^*sz*^: *X*^*sz*^ was normalized to *X*^*base*^ using the RobustScaler algorithm. After generating windowed *NDD* values, we then applied the same post-processing smoothing steps as described above. We then generated binary seizure detections over a sweep of thresholds to calculate AUROC.

We also benchmark *NDD* predictions against three previously published and validated seizure detection models. The first two benchmark models were the RAMSES model, which had been developed on a subset of our ICU cohort in a previous publication^36^ and the kaggle seizure detection algorithm, which was developed and tested for a seizure detection competition^37^. Both of these algorithms require pretraining, and so their predictions were evaluated in a cross-validation paradigm. We also benchmarked *NDD* against SPaRCNet, a state of the art pretrained scalp EEG model^38^.

### L. Statistical Methods

When comparing distributions at the patient level (e.g., diffuse vs. focal epilepsies, scalp EEG model performance) we use non-parametric Mann-Whitney U tests to avoid making assumptions about the distributions. When making comparisons at the seizure level we use a linear mixed-effects model to account for pooled variance within patients. Throughout the manuscript we primarily leverage non-parametric statistical tests to avoid any assumptions about the normality of statistical distributions. For all validation experiments and analyses, tests were performed at the seizure level unless otherwise noted. Within experiments, we correct for multiple comparisons and reduce the rate of false positives using the Benjamini-Hochberg procedure^75^. All comparisons were two-tailed unless otherwise noted. We considered any test with a p value below 0.10 to be a trending effect with an *α* of 0.05 to assess statistical significance.

## Notes

### Competing Interest Statement

The authors have declared no competing interest.

### Funding Statement

This work was funded by National Institute of Neurologic Disorders and Stroke (DJZ: 5T32NS091008; RTS: R01MH112847, R01NS112274; KAD: R01NS116504, BL: DP1NS122038, R01NS125137; EC: K23NS12140101A1), the National Science Foundation (WKSO: NSF GRF DGE-1845298), the Burroughs Welcome Fund (EC), the Small Lake Foundation, Neil and Barbara Smit, and Bonnie and Jonathan Rothberg and Family (BL).

### Author Declarations

The collection and retrospective analysis of all data described in the manuscript was given ethical approval by the IRB of University of Pennsylvania.

