## Supplementary Material for "Unsupervised seizure annotation and detection with neural dynamic divergence"

### A. Model performance metrics.

**Table Supplementary Table 1.** Model Performance Summary: Mean ± 95% CI

| Stage | Metric | ABSSLP | HFER | IMPRINT | Interrater | LiNDDA | NDD | WVNT |
|---|---|---|---|---|---|---|---|---|
| test | opt. onset $\phi$ | 0.32 ± 0.09 | 0.29 ± 0.07 | 0.56 ± 0.08 | **0.64 ± 0.07** | **0.58 ± 0.09** | 0.57 ± 0.10 | 0.55 ± 0.09 |
| test | opt. spread $\phi$ | 0.45 ± 0.08 | 0.34 ± 0.07 | 0.59 ± 0.06 | 0.63 ± 0.06 | **0.69 ± 0.07** | 0.66 ± 0.08 | 0.62 ± 0.07 |
| test | onset auroc | 0.62 ± 0.09 | 0.68 ± 0.07 | **0.88 ± 0.06** | – | **0.88 ± 0.06** | 0.81 ± 0.08 | 0.80 ± 0.08 |
| test | onset auprc raw | 0.20 ± 0.09 | 0.15 ± 0.07 | 0.39 ± 0.09 | – | **0.47 ± 0.11** | **0.47 ± 0.11** | 0.43 ± 0.11 |
| test | pre. onset $\phi$ | 0.10 ± 0.05 | 0.10 ± 0.04 | 0.30 ± 0.08 | **0.64 ± 0.07** | **0.35 ± 0.09** | **0.35 ± 0.08** | 0.31 ± 0.08 |
| test | spread auroc | 0.73 ± 0.08 | 0.67 ± 0.07 | 0.89 ± 0.05 | – | **0.90 ± 0.05** | 0.88 ± 0.05 | 0.88 ± 0.05 |
| test | spread auprc raw | 0.40 ± 0.09 | 0.28 ± 0.08 | 0.56 ± 0.08 | – | **0.65 ± 0.08** | 0.63 ± 0.09 | 0.60 ± 0.08 |
| test | pre. spread $\phi$ | 0.22 ± 0.06 | 0.13 ± 0.04 | 0.38 ± 0.06 | **0.63 ± 0.06** | 0.44 ± 0.07 | **0.45 ± 0.07** | 0.39 ± 0.06 |
| validation | spread rank | 28.26 ± 1.24 | 33.91 ± 1.23 | 21.66 ± 1.16 | – | **14.98 ± 1.14** | 14.91 ± 1.11 | 18.58 ± 1.17 |
| validation | auroc | 0.67 ± 0.02 | 0.67 ± 0.01 | 0.80 ± 0.01 | – | **0.87 ± 0.01** | 0.85 ± 0.01 | 0.82 ± 0.01 |
| validation | auprc | 0.28 ± 0.02 | 0.27 ± 0.02 | 0.36 ± 0.02 | – | **0.54 ± 0.02** | 0.53 ± 0.02 | 0.48 ± 0.02 |
| validation | recruitment latency | 16.53 ± 2.07 | 25.44 ± 2.61 | 11.97 ± 1.73 | – | **9.83 ± 1.52** | 10.66 ± 1.52 | 10.02 ± 1.50 |
| validation | f1 | 0.22 ± 0.01 | 0.18 ± 0.01 | 0.29 ± 0.01 | – | **0.42 ± 0.02** | **0.42 ± 0.01** | 0.36 ± 0.01 |
| validation | $\phi$ | 0.17 ± 0.01 | 0.12 ± 0.01 | 0.27 ± 0.01 | – | **0.42 ± 0.02** | 0.41 ± 0.02 | 0.34 ± 0.02 |
| validation | precision | 0.16 ± 0.01 | 0.14 ± 0.01 | 0.26 ± 0.02 | – | **0.40 ± 0.02** | **0.40 ± 0.02** | 0.32 ± 0.02 |
| validation | sensitivity | 0.57 ± 0.03 | 0.55 ± 0.03 | 0.62 ± 0.02 | – | **0.69 ± 0.02** | 0.67 ± 0.02 | 0.68 ± 0.02 |
| validation | specificity | 0.71 ± 0.02 | 0.64 ± 0.02 | 0.77 ± 0.01 | – | 0.85 ± 0.01 | **0.86 ± 0.01** | 0.80 ± 0.01 |

### B. Model comparison statistics.

**Table Supplementary Table 2.** Model Performance Comparison Statistics

| Stage | Model 1 | Model 2 | Metric | p-value | Intercept | Rank | FDR-corrected p |
|---|---|---|---|---|---|---|---|
| test | LiNDDA | ABSSLP | max_onset_phi | 1.05e-04 | 0.22 | 4 | 2.64e-04 |
| test | LiNDDA | HFER | max_onset_phi | 1.45e-07 | 0.27 | 1 | 1.45e-06 |
| test | LiNDDA | IMPRINT | max_onset_phi | 0.789 | 0.01 | 9 | 0.876 |
| test | LiNDDA | Interrater | max_onset_phi | 0.137 | -0.07 | 6 | 0.229 |
| test | LiNDDA | WVNT | max_onset_phi | 0.473 | 0.02 | 7 | 0.676 |
| test | NDD | ABSSLP | max_onset_phi | 9.03e-05 | 0.22 | 3 | 2.64e-04 |
| test | NDD | HFER | max_onset_phi | 1.03e-06 | 0.26 | 2 | 5.13e-06 |
| test | NDD | IMPRINT | max_onset_phi | 0.949 | 0.00 | 10 | 0.949 |
| test | NDD | Interrater | max_onset_phi | 0.068 | -0.09 | 5 | 0.137 |
| test | NDD | WVNT | max_onset_phi | 0.622 | 0.01 | 8 | 0.778 |
| test | LiNDDA | ABSSLP | max_spread_phi | 2.25e-07 | 0.21 | 3 | 7.49e-07 |
| test | LiNDDA | HFER | max_spread_phi | 1.38e-11 | 0.33 | 1 | 1.38e-10 |
| test | LiNDDA | IMPRINT | max_spread_phi | 0.002 | 0.09 | 5 | 0.004 |
| test | LiNDDA | Interrater | max_spread_phi | 0.049 | 0.06 | 7 | 0.070 |
| test | LiNDDA | WVNT | max_spread_phi | 0.004 | 0.07 | 6 | 0.007 |
| test | NDD | ABSSLP | max_spread_phi | 2.70e-05 | 0.18 | 4 | 6.76e-05 |
| test | NDD | HFER | max_spread_phi | 4.18e-08 | 0.29 | 2 | 2.09e-07 |
| test | NDD | IMPRINT | max_spread_phi | 0.097 | 0.06 | 8 | 0.110 |
| test | NDD | Interrater | max_spread_phi | 0.522 | 0.02 | 10 | 0.522 |
| test | NDD | WVNT | max_spread_phi | 0.099 | 0.04 | 9 | 0.110 |
| test | LiNDDA | ABSSLP | onset_auc | 1.32e-11 | 0.26 | 1 | 1.06e-10 |
| test | LiNDDA | HFER | onset_auc | 3.62e-07 | 0.20 | 2 | 1.45e-06 |
| test | LiNDDA | IMPRINT | onset_auc | 0.692 | -0.01 | 8 | 0.692 |
| test | LiNDDA | WVNT | onset_auc | 0.008 | 0.08 | 4 | 0.014 |
| test | NDD | ABSSLP | onset_auc | 2.02e-06 | 0.19 | 3 | 5.37e-06 |
| test | NDD | HFER | onset_auc | 0.009 | 0.13 | 5 | 0.014 |
| test | NDD | IMPRINT | onset_auc | 0.047 | -0.07 | 6 | 0.063 |
| test | NDD | WVNT | onset_auc | 0.597 | 0.01 | 7 | 0.683 |
| test | LiNDDA | ABSSLP | onset_auprc_normalized | 0.005 | 0.23 | 4 | 0.009 |
| test | LiNDDA | HFER | onset_auprc_normalized | 1.01e-06 | 0.31 | 1 | 8.05e-06 |
| test | LiNDDA | IMPRINT | onset_auprc_normalized | 0.323 | 0.06 | 6 | 0.431 |
| test | LiNDDA | WVNT | onset_auprc_normalized | 0.767 | 0.02 | 7 | 0.771 |
| test | NDD | ABSSLP | onset_auprc_normalized | 0.002 | 0.24 | 3 | 0.004 |
| test | NDD | HFER | onset_auprc_normalized | 3.05e-06 | 0.31 | 2 | 1.22e-05 |
| test | NDD | IMPRINT | onset_auprc_normalized | 0.286 | 0.06 | 5 | 0.431 |
| test | NDD | WVNT | onset_auprc_normalized | 0.771 | 0.02 | 8 | 0.771 |
| test | LiNDDA | ABSSLP | onset_phi_at_learned_f1_plateau_median | 1.22e-04 | 0.22 | 2 | 3.96e-04 |
| test | LiNDDA | HFER | onset_phi_at_learned_f1_plateau_median | 9.83e-05 | 0.22 | 1 | 3.96e-04 |
| test | LiNDDA | IMPRINT | onset_phi_at_learned_f1_plateau_median | 0.288 | 0.05 | 5 | 0.363 |
| test | LiNDDA | WVNT | onset_phi_at_learned_f1_plateau_median | 0.476 | 0.03 | 8 | 0.476 |
| test | NDD | ABSSLP | onset_phi_at_learned_f1_plateau_median | 1.82e-04 | 0.21 | 3 | 3.96e-04 |
| test | NDD | HFER | onset_phi_at_learned_f1_plateau_median | 1.98e-04 | 0.21 | 4 | 3.96e-04 |
| test | NDD | IMPRINT | onset_phi_at_learned_f1_plateau_median | 0.291 | 0.05 | 6 | 0.363 |
| test | NDD | WVNT | onset_phi_at_learned_f1_plateau_median | 0.317 | 0.03 | 7 | 0.363 |
| test | LiNDDA | ABSSLP | spread_auc | 3.27e-05 | 0.16 | 2 | 1.31e-04 |
| test | LiNDDA | HFER | spread_auc | 1.19e-07 | 0.22 | 1 | 9.49e-07 |

Continued on next page

Table Supplementary Table 2 – continued from previous page

| Stage | Model 1 | Model 2 | Metric | p-value | Intercept | Rank | FDR-corrected p |
|---|---|---|---|---|---|---|---|
| test | LiNDDA | IMPRINT | spread_auc | 0.543 | 0.01 | 6 | 0.723 |
| test | LiNDDA | WVNT | spread_auc | 0.368 | 0.02 | 5 | 0.588 |
| test | NDD | ABSSLP | spread_auc | 0.001 | 0.13 | 4 | 0.003 |
| test | NDD | HFER | spread_auc | 1.18e-04 | 0.20 | 3 | 3.14e-04 |
| test | NDD | IMPRINT | spread_auc | 0.652 | -0.01 | 7 | 0.746 |
| test | NDD | WVNT | spread_auc | 0.826 | 0.00 | 8 | 0.826 |
| test | LiNDDA | ABSSLP | spread_auprc_normalized | 1.33e-04 | 0.24 | 3 | 3.56e-04 |
| test | LiNDDA | HFER | spread_auprc_normalized | 6.39e-10 | 0.38 | 1 | 5.11e-09 |
| test | LiNDDA | IMPRINT | spread_auprc_normalized | 0.116 | 0.09 | 6 | 0.155 |
| test | LiNDDA | WVNT | spread_auprc_normalized | 0.099 | 0.07 | 5 | 0.155 |
| test | NDD | ABSSLP | spread_auprc_normalized | 3.80e-04 | 0.21 | 4 | 7.61e-04 |
| test | NDD | HFER | spread_auprc_normalized | 5.80e-07 | 0.35 | 2 | 2.32e-06 |
| test | NDD | IMPRINT | spread_auprc_normalized | 0.311 | 0.06 | 7 | 0.327 |
| test | NDD | WVNT | spread_auprc_normalized | 0.327 | 0.03 | 8 | 0.327 |
| test | LiNDDA | ABSSLP | spread_phi_at_learned_f1_plateau_median | 3.75e-04 | 0.19 | 4 | 7.50e-04 |
| test | LiNDDA | HFER | spread_phi_at_learned_f1_plateau_median | 4.74e-11 | 0.31 | 2 | 1.90e-10 |
| test | LiNDDA | IMPRINT | spread_phi_at_learned_f1_plateau_median | 0.334 | 0.04 | 8 | 0.334 |
| test | LiNDDA | WVNT | spread_phi_at_learned_f1_plateau_median | 0.084 | 0.04 | 6 | 0.112 |
| test | NDD | ABSSLP | spread_phi_at_learned_f1_plateau_median | 3.08e-05 | 0.21 | 3 | 8.20e-05 |
| test | NDD | HFER | spread_phi_at_learned_f1_plateau_median | 2.81e-11 | 0.31 | 1 | 1.90e-10 |
| test | NDD | IMPRINT | spread_phi_at_learned_f1_plateau_median | 0.172 | 0.06 | 7 | 0.197 |
| test | NDD | WVNT | spread_phi_at_learned_f1_plateau_median | 0.001 | 0.06 | 5 | 0.002 |
| validation | LiNDDA | ABSSLP | adj_med_soz_spread_rank_pct | 1.17e-19 | -12.26 | 1 | 9.39e-19 |
| validation | LiNDDA | HFER | adj_med_soz_spread_rank_pct | 9.15e-19 | -16.62 | 3 | 2.44e-18 |
| validation | LiNDDA | IMPRINT | adj_med_soz_spread_rank_pct | 4.41e-06 | -5.76 | 5 | 6.87e-06 |
| validation | LiNDDA | WVNT | adj_med_soz_spread_rank_pct | 0.008 | -1.84 | 8 | 0.008 |
| validation | NDD | ABSSLP | adj_med_soz_spread_rank_pct | 2.13e-17 | -12.55 | 4 | 4.27e-17 |
| validation | NDD | HFER | adj_med_soz_spread_rank_pct | 6.71e-19 | -17.06 | 2 | 2.44e-18 |
| validation | NDD | IMPRINT | adj_med_soz_spread_rank_pct | 5.15e-06 | -6.08 | 6 | 6.87e-06 |
| validation | NDD | WVNT | adj_med_soz_spread_rank_pct | 8.57e-05 | -2.14 | 7 | 9.80e-05 |
| validation | LiNDDA | ABSSLP | auc | 7.96e-26 | 0.20 | 1 | 6.37e-25 |
| validation | LiNDDA | HFER | auc | 1.25e-15 | 0.17 | 3 | 3.34e-15 |
| validation | LiNDDA | IMPRINT | auc | 2.57e-05 | 0.05 | 7 | 2.94e-05 |
| validation | LiNDDA | WVNT | auc | 1.20e-09 | 0.05 | 5 | 1.92e-09 |
| validation | NDD | ABSSLP | auc | 5.23e-18 | 0.18 | 2 | 2.09e-17 |
| validation | NDD | HFER | auc | 3.47e-10 | 0.14 | 4 | 6.94e-10 |
| validation | NDD | IMPRINT | auc | 0.031 | 0.03 | 8 | 0.031 |
| validation | NDD | WVNT | auc | 2.06e-06 | 0.02 | 6 | 2.74e-06 |
| validation | LiNDDA | ABSSLP | avg_soz_recruitment_latency | 1.33e-07 | -7.78 | 3 | 3.54e-07 |
| validation | LiNDDA | HFER | avg_soz_recruitment_latency | 6.11e-12 | -23.36 | 1 | 4.89e-11 |
| validation | LiNDDA | IMPRINT | avg_soz_recruitment_latency | 0.007 | -3.14 | 5 | 0.012 |
| validation | LiNDDA | WVNT | avg_soz_recruitment_latency | 0.351 | -0.44 | 8 | 0.351 |
| validation | NDD | ABSSLP | avg_soz_recruitment_latency | 1.06e-05 | -6.50 | 4 | 2.13e-05 |
| validation | NDD | HFER | avg_soz_recruitment_latency | 1.09e-10 | -22.00 | 2 | 4.35e-10 |
| validation | NDD | IMPRINT | avg_soz_recruitment_latency | 0.065 | -2.07 | 6 | 0.086 |
| validation | NDD | WVNT | avg_soz_recruitment_latency | 0.091 | 0.82 | 7 | 0.104 |
| validation | LiNDDA | ABSSLP | f1 | 3.99e-23 | 0.18 | 1 | 3.19e-22 |
| validation | LiNDDA | HFER | f1 | 8.92e-23 | 0.21 | 2 | 3.57e-22 |
| validation | LiNDDA | IMPRINT | f1 | 1.20e-14 | 0.11 | 5 | 1.92e-14 |
| validation | LiNDDA | WVNT | f1 | 5.22e-08 | 0.05 | 8 | 5.22e-08 |
| validation | NDD | ABSSLP | f1 | 4.06e-21 | 0.17 | 4 | 8.13e-21 |
| validation | NDD | HFER | f1 | 2.56e-21 | 0.21 | 3 | 6.83e-21 |
| validation | NDD | IMPRINT | f1 | 4.98e-13 | 0.10 | 6 | 6.64e-13 |
| validation | NDD | WVNT | f1 | 2.88e-08 | 0.05 | 7 | 3.29e-08 |
| validation | LiNDDA | ABSSLP | phi | 6.43e-27 | 0.22 | 1 | 5.15e-26 |
| validation | LiNDDA | HFER | phi | 2.11e-26 | 0.26 | 2 | 8.42e-26 |
| validation | LiNDDA | IMPRINT | phi | 4.15e-16 | 0.12 | 5 | 6.64e-16 |
| validation | LiNDDA | WVNT | phi | 1.68e-10 | 0.06 | 8 | 1.68e-10 |
| validation | NDD | ABSSLP | phi | 7.14e-24 | 0.21 | 4 | 1.43e-23 |
| validation | NDD | HFER | phi | 3.50e-24 | 0.25 | 3 | 9.33e-24 |
| validation | NDD | IMPRINT | phi | 3.87e-14 | 0.11 | 6 | 5.16e-14 |
| validation | NDD | WVNT | phi | 2.54e-11 | 0.05 | 7 | 2.90e-11 |
| validation | LiNDDA | ABSSLP | precision | 8.46e-25 | 0.22 | 1 | 6.77e-24 |
| validation | LiNDDA | HFER | precision | 3.02e-20 | 0.24 | 3 | 8.05e-20 |
| validation | LiNDDA | IMPRINT | precision | 1.07e-16 | 0.13 | 5 | 1.71e-16 |
| validation | LiNDDA | WVNT | precision | 1.17e-10 | 0.08 | 8 | 1.17e-10 |
| validation | NDD | ABSSLP | precision | 2.96e-21 | 0.21 | 2 | 1.18e-20 |
| validation | NDD | HFER | precision | 6.20e-18 | 0.22 | 4 | 1.24e-17 |
| validation | NDD | IMPRINT | precision | 1.11e-14 | 0.11 | 6 | 1.48e-14 |
| validation | NDD | WVNT | precision | 6.73e-12 | 0.07 | 7 | 7.69e-12 |
| validation | LiNDDA | ABSSLP | sensitivity | 8.82e-08 | 0.13 | 1 | 7.05e-07 |
| validation | LiNDDA | HFER | sensitivity | 8.66e-07 | 0.18 | 2 | 3.46e-06 |
| validation | LiNDDA | IMPRINT | sensitivity | 0.029 | 0.05 | 6 | 0.039 |
| validation | LiNDDA | WVNT | sensitivity | 0.013 | 0.03 | 5 | 0.020 |
| validation | NDD | ABSSLP | sensitivity | 1.49e-04 | 0.11 | 4 | 2.98e-04 |
| validation | NDD | HFER | sensitivity | 9.80e-05 | 0.16 | 3 | 2.61e-04 |
| validation | NDD | IMPRINT | sensitivity | 0.274 | 0.03 | 7 | 0.313 |
| validation | NDD | WVNT | sensitivity | 0.598 | 0.01 | 8 | 0.598 |
| validation | LiNDDA | ABSSLP | specificity | 2.21e-13 | 0.10 | 2 | 8.86e-13 |

Continued on next page

Table Supplementary Table 2 – continued from previous page

| Stage | Model 1 | Model 2 | Metric | p-value | Intercept | Rank | FDR-corrected p |
|---|---|---|---|---|---|---|---|
| validation | LiNDDA | HFER | specificity | 8.11e-07 | 0.12 | 7 | 9.27e-07 |
| validation | LiNDDA | IMPRINT | specificity | 5.47e-09 | 0.06 | 6 | 7.29e-09 |
| validation | LiNDDA | WVNT | specificity | 0.049 | 0.02 | 8 | 0.049 |
| validation | NDD | ABSSLP | specificity | 3.74e-21 | 0.12 | 1 | 2.99e-20 |
| validation | NDD | HFER | specificity | 4.22e-10 | 0.14 | 5 | 6.76e-10 |
| validation | NDD | IMPRINT | specificity | 9.48e-13 | 0.08 | 3 | 2.40e-12 |
| validation | NDD | WVNT | specificity | 1.20e-12 | 0.04 | 4 | 2.40e-12 |

**C. Optimizing hyperparameters for iEEG seizure annotation.** The $NDD$ LSTM was set to have a sequence length of $s = 12$ and a hidden state dimension of 10 [76], and the model was trained using batched gradient descent over 10 epochs, with batch size, $\mathcal{B}$, set to $\mathcal{B} = [\frac{L-s}{2}]$. These hyperparameters enable a shared low-dimensional latent space across channels capable of capturing seizure dynamics [18], spatial correlations and simplifying the dynamics relative to single-channel models [63]. The $LiNDDA$ model was trained using batched gradient descent over a maximum of 50 epochs and batch size, $\mathcal{B} = 512$, and early stopping after loss did not improve beyond a cutoff of 1e-04 for two epochs on a causal held out validation split of 10% of $X^{base}$. All models were trained with the Adam optimizer (default PyTorch settings) and a learning rate of 0.01 in a teacher-forcing paradigm, only forecasting one time step in the future. To identify the optimal autoregressive order for the $LiNDDA$ architecture, we leveraged the un-thresholded validation data and optimized model order based on AUROC and AUPRC for seizure onset localization. To narrow the hyperparameter space, we set the forecasting horizon to $n - 1$ steps for each order $n$, allowing us to explore the tradeoff between model complexity and dynamic interrogation. Based on both performance metrics, we selected an order of 3 (i.e., 3 time steps of input) for the $LiNDDA$ model (Figure S1). Although the model was trained using next-time-step prediction error, we generated $s - 1$ time steps of predictions (in this case, 2) when measuring dynamic divergence, more thoroughly probing the learned dynamics encoded in the surrogate brain model. The convex performance curve suggests that even lower-order models contained sufficient parameters to capture multivariate neural dynamics, likely reflecting the relatively low intrinsic dimensionality of neural activity recorded from large electrode arrays [77].

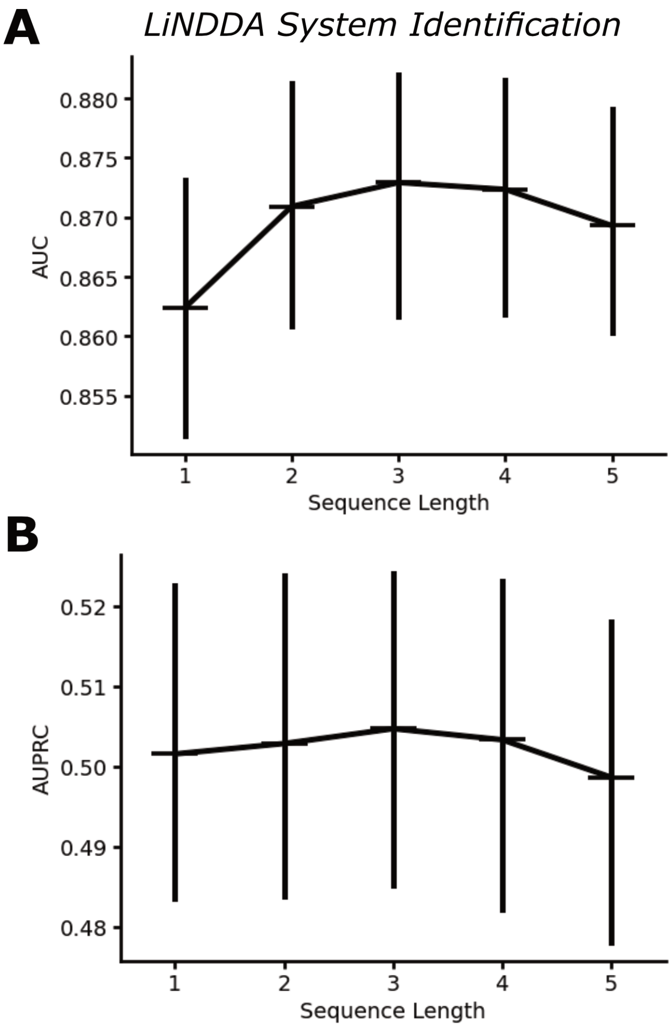


**Fig. S1. System identification for LiNDDA Models**:
**A**) AUROC and **B**) AUPRC distributions show performance of $LiNDDA$ models with different orders of sequence length. Models forecasted $s - 1$ time steps into the future when running inference.

**D. Seizure semiology comparison statistics.**

**Table Supplementary Table 3.** Seizure Spread and Semiological Severity

| Seizure Semiology 1 | Seizure Semiology 2 | p-value | Rank | FDR-corrected p |
|---|---|---|---|---|
| Subclinical | FAS | 1.94e-04 | 2 | 2.91e-04 |
| FAS | FIAS | 0.560 | 3 | 0.560 |
| FIAS | FBTCS | 6.78E-09 | 1 | 2.03E-08 |

**E. Optimizing $LiNDDA$ hyperparameters for Scalp EEG seizure detection.** When deploying the $NDD$ model with the $LiNDDA$ backbone on scalp EEG datasets, we tested two different sequence and forecast length hyperparameters on the publically available validation dataset: sequence length/forecast length 5/4 and 3/2 Supplementary Table 4. The AUROC for continuous seizure detection was highest in the $LiNDDA-54$ model, which we then deploy on the test dataset and compare to other benchmark models.

**F. Scalp EEG Seizure Detection Performance Metrics.**

**Table Supplementary Table 4.** Scalp EEG Model Performance: AUROC Metrics

| Analysis Set | Model | Mean AUROC | Median AUROC | n |
|---|---|---|---|---|
| Validation Set | Kaggle | 0.74 ± 0.04 | 0.77 ± 0.06 | 36 |
| Validation Set | LiNDDA-32 | 0.78 ± 0.06 | 0.82 ± 0.06 | 36 |
| Validation Set | LiNDDA-54 | **0.80 ± 0.05** | **0.84 ± 0.07** | 36 |
| Validation Set | NDD | **0.80 ± 0.06** | **0.84 ± 0.07** | 36 |
| Validation Set | RAMSES | 0.68 ± 0.05 | 0.69 ± 0.07 | 36 |
| Validation Set | SparcNet | 0.78 ± 0.05 | 0.82 ± 0.08 | 36 |
| Test Set | Kaggle | 0.64 ± 0.11 | 0.65 ± 0.11 | 14 |
| Test Set | LiNDDA-32 | 0.68 ± 0.11 | 0.64 ± 0.14 | 14 |
| Test Set | LiNDDA-54 | 0.68 ± 0.11 | **0.66 ± 0.14** | 14 |
| Test Set | NDD | **0.69 ± 0.10** | 0.63 ± 0.12 | 14 |
| Test Set | RAMSES | 0.51 ± 0.08 | 0.50 ± 0.08 | 14 |
| Test Set | SparcNet | 0.58 ± 0.10 | 0.61 ± 0.09 | 14 |
| Combined | Kaggle | 0.71 ± 0.04 | 0.75 ± 0.06 | 50 |
| Combined | LiNDDA-32 | 0.75 ± 0.05 | 0.80 ± 0.07 | 50 |
| Combined | LiNDDA-54 | **0.77 ± 0.05** | 0.79 ± 0.10 | 50 |
| Combined | NDD | **0.77 ± 0.05** | **0.81 ± 0.07** | 50 |
| Combined | RAMSES | 0.63 ± 0.05 | 0.63 ± 0.05 | 50 |
| Combined | SparcNet | 0.72 ± 0.05 | 0.73 ± 0.10 | 50 |

**G. Scalp EEG Seizure Detection Comparison Statistics.**

**Table Supplementary Table 5.** Scalp EEG Model Statistical Comparisons

| Analysis Set | Model | Benchmark | Mean Diff | p-value | Rank | FDR-corrected p | n |
|---|---|---|---|---|---|---|---|
| Combined | LiNDDA-54 | RAMSES | 0.13 | 2.84e-07 | 2 | 8.52e-07 | 50 |
| Combined | LiNDDA-54 | Kaggle | 0.05 | 0.018 | 4 | 0.026 | 50 |
| Combined | LiNDDA-54 | SparcNet | 0.04 | 0.099 | 6 | 0.099 | 50 |
| Combined | NDD | RAMSES | 0.14 | 2.47e-07 | 1 | 8.52e-07 | 50 |
| Combined | NDD | Kaggle | 0.06 | 0.006 | 3 | 0.013 | 50 |
| Combined | NDD | SparcNet | 0.05 | 0.055 | 5 | 0.067 | 50 |

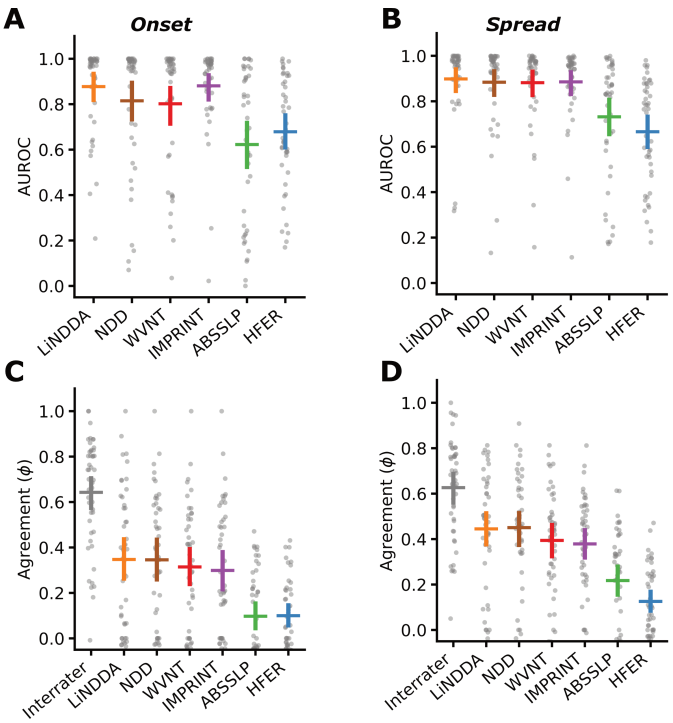


**Fig. S2. Additional performance metrics on gold-standard annotated seizures**: The AUROC for benchmark and $NDD$ models classifying (**A**) onset and (**B**) spread activity from non-seiziing channels at seizure onset and after 10 seconds of seizure spread on the gold-standard consensus dataset. On the same set of seizures, we also compare model agreement at the fixed, pre-trained threshold (Figure 3H) and see that the $NDD$ models are state of the art for both (**C**) onset and (**D**) spread annotation (Supplementary Table 1).

#### H. Model Performance On Consensus Annotations.

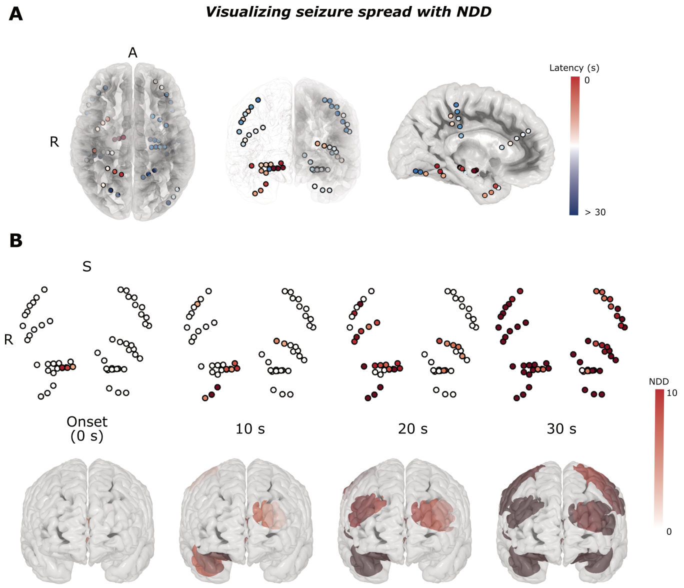


**Fig. S3. Integration of $NDD$ with seizure visualization software**: Here, we integrate NDD seizure annotations with the RAVE visualization software[56]. **A**) We show electrodes co-registered to an MNI brain shaded by seizure spread latency highlighting seizure onset in the right hippocampus and rapidly spreading to the contralateral frontal lobe. **B**) We also show how NDD can be used directly without thresholding to visualize the spread of abnormality of seizure activity through implanted electrodes (top) and projected onto the cortical surface (bottom).

**I. Seizure Spread Visualization.**
